# Predicting the impact of COVID-19 interruptions on transmission of gambiense human African trypanosomiasis in two health zones of the Democratic Republic of Congo

**DOI:** 10.1101/2020.10.26.20219485

**Authors:** Maryam Aliee, Soledad Castaño, Christopher N Davis, Swati Patel, Erick Mwamba Miaka, Simon EF Spencer, Matt J Keeling, Nakul Chitnis, Kat S Rock

**Affiliations:** Mathematics Institute, University of Warwick, Coventry, CV4 7AL, United Kingdom; Zeeman Institute: SBIDER, University of Warwick, Coventry, CV4 7AL, United Kingdom; Department of Epidemiology and Public Health, Swiss Tropical and Public Health Institute, Basel, Switzerland; University of Basel, Basel, Switzerland; Department of Statistics, The University of Warwick, Coventry, CV4 7AL, United Kingdom; Programme National de Lutte contre la Trypanosomiase Humaine Africaine, Kinshasa, the Democratic Republic of the Congo; School of Life Sciences, University of Warwick, Coventry, CV4 7AL, United Kingdom

**Keywords:** Gambiense human African trypanosomiasis (gHAT), modelling, elimination of transmission, COVID-19, mitigation

## Abstract

Many control programmes against neglected tropical diseases have been interrupted due to COVID-19 pandemic, including those that rely on active case finding. In this study we focus on gambiense human African trypanosomiasis (gHAT), where active screening was suspended in the Democratic Republic of Congo (DRC) due to the pandemic. We use two independent mathematical models to predict the impact of COVID-19 interruptions on transmission and reporting, and the achievement of 2030 elimination of transmission (EOT) goal for gHAT in two moderate-risk regions of DRC. We consider different interruption scenarios, including reduced passive surveillance in fixed health facilities, and whether this suspension lasts until the end of 2020 or 2021. Our models predict an increase in the number of new infections in the interruption period only if both active screening and passive surveillance were suspended, and with slowed reduction - but no increase - if passive surveillance remains fully functional. In all scenarios, the EOT may be slightly pushed back if no mitigation such as increased screening coverage is put in place. However, we emphasise that the biggest challenge will remain in the higher prevalence regions where EOT is already predicted to be behind schedule without interruptions unless interventions are bolstered.

## Introduction

The threat posed by the 2019 coronavirus disease (COVID-19) pandemic is not limited to the direct consequences of the disease itself. In addition to the economic burden that many countries are facing due to lockdowns, the public health measures initiated to suppress transmission of the coronavirus, SARS-CoV-2, may have an additional impact on the ability to control other infectious diseases. A disruption in usual activities of health services has the potential to lead to an increased loss of life as surveillance and access to diagnostics and treatment is more limited (Wang, 2020). These challenges to public health systems will affect Africa disproportionately, with COVID-19 providing an additional burden to many already fragile health systems (Makoni, 2020); and previous modelling studies have suggested that COVID-19 related interruptions to malaria control could lead to substantially higher cases and deaths in Africa in the near future (Sherrard-Smith 2020, Hogan 2020).

*Gambiense* human African trypanosomiasis (gHAT) is a vector-borne disease of Central and West Africa, which is typically fatal when left untreated, for which the delivery of interventions has already been impacted by the COVID-19 pandemic. A primary intervention to control gHAT is the use of active screening (AS), where at-risk populations in hard-to-reach locations are tested using serological diagnostics followed by subsequent case confirmation and treatment. In April 2020, the World Health Organization (WHO) recommended that active case-finding activities and mass treatment campaigns for neglected tropical diseases should be postponed until further notice (WHO, 2020). The lack of AS results in a reliance on the passive healthcare system and individuals self-presenting to health centres following onset of symptoms. This will likely lead to diagnosis at a later stage of this lethal disease, with additional uncertainty about whether the pandemic will also lead to a fear of travelling to health centres by patients, and a reduced priority of the disease for diagnosis in the health centres due to COVID-19 testing (Camara, 2017, Chanda-Kapata, 2020).

For gHAT, interruptions in AS in the past have led to increased infections in subsequent years. The 2014–2015 Ebola outbreak in Guinea, which completely interrupted AS and resulted in “partial” passive surveillance, coincided with a fall in reported gHAT incidence, which was likely due to poor surveillance rather than a true reduction in infection numbers (Camara, 2017). We can also learn from other interruptions of AS: a period of no active screening in 2007–2008 in Mandoul, Chad, also led to increased passive case detections in 2008, with modelling suggesting there was an increase in transmission (Mahamat, 2017). A challenge for controlling intensified disease management neglected tropical diseases (IDM NTDs) is the strong link between control activities (i.e. screening) and surveillance through case reporting, so a reduction in reported cases can be indicative of either success (if surveillance is strong) or reduced invention effort.

There is a risk that the interruption of gHAT interventions will impact the goal of elimination of transmission (EOT) of gHAT by 2030, which has been set by the WHO (Franco, 2020). While modelling suggests that it is already unlikely that all areas will achieve this goal (Huang 2020, NTD-gHAT 2020), there is now the potential for further delays. To quantify the length of a delay in time to reach EOT caused by COVID-19 interruptions, and the corresponding increase in transmission, cases and possible deaths, we use two independent stochastic models for gHAT infection in two distinct transmission settings in the Democratic Republic of Congo (DRC), the country which has the highest case burden.

## Methods

In this work, we focus on administrative regions in Kwilu province (within the former Bandundu province) which was the highest endemicity province in the country in 2016. We selected two different health zones within Kwilu, Bagata and Mosango, both classified as moderate-risk in 2016 (1-10 reported cases per 10,000 people per year (Franco et al, 2020)), although Bagata has had higher case reporting (see SI Fig 2,4,5). Both health zones are similar in population size (estimated as 121,433 for Mosango and 165,990 for Bagata in 2015 (OCHA)). The health zones have had regular active screening with Bagata generally observing higher coverage: in Bagata the mean coverage during 2014-2018 was approximately 35%, and the maximum was around 45%; in Mosango the mean coverage during 2014-2018 was around 33%, and the maximum was 60%. We chose regions in which vector control has not yet been implemented in order to better study the significance of interruptions of screening.

**Fig 1.**
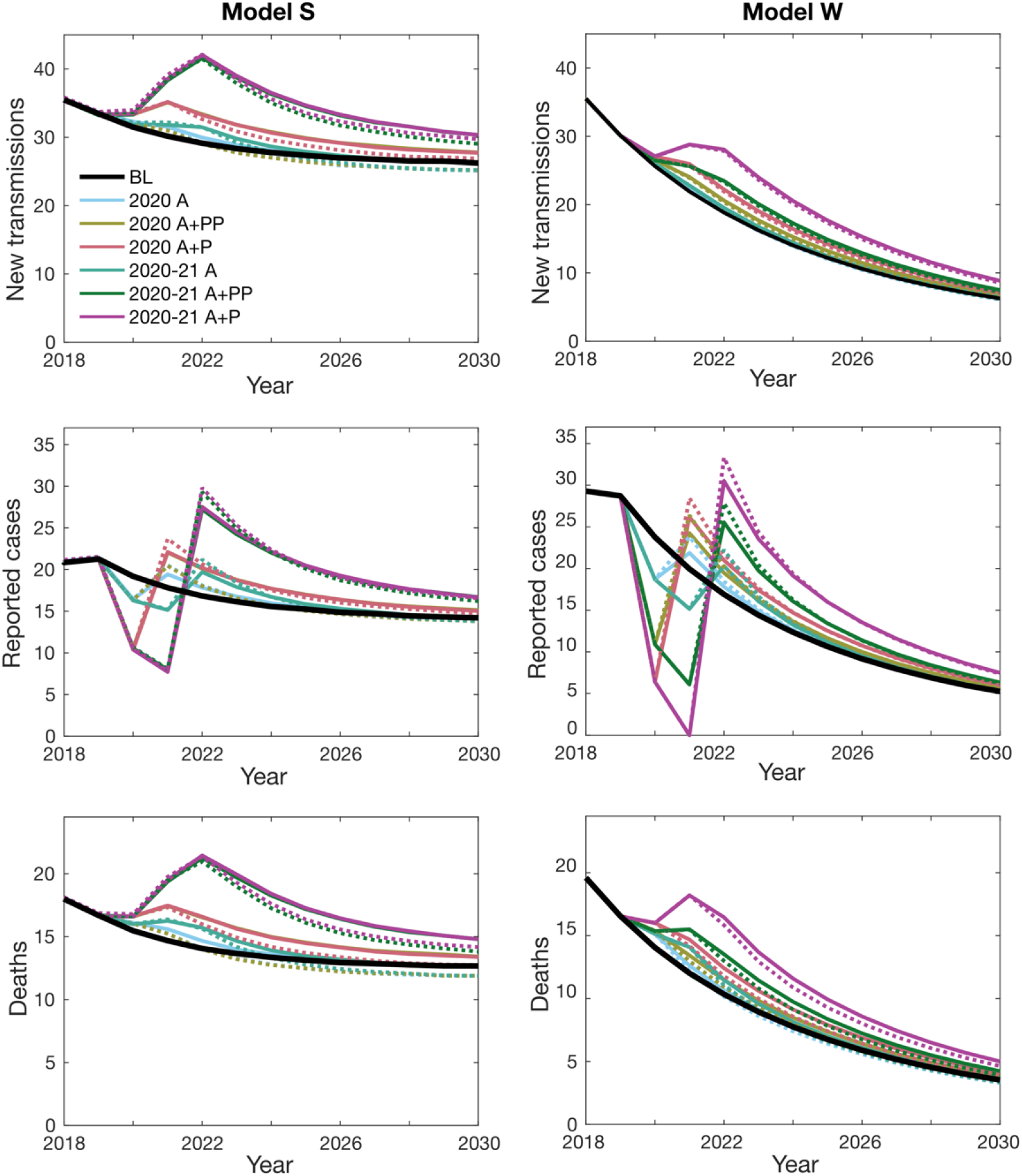
Predicted gHAT infections in the health zone Bagata based on different COVID-19 interruption scenarios. The graphs show the expected number of new transmissions, reported cases, and number of deaths caused by the disease (mean values), for Model S (left hand side) and Model W (right hand side). The baseline (BL) is shown as a black solid line. Individual interruption scenarios without mitigation are depicted by solid lines with different colours as indicated in the first panel. The corresponding mitigation scenarios are shown with dashed lines using comparable colours.

**Fig 2.**
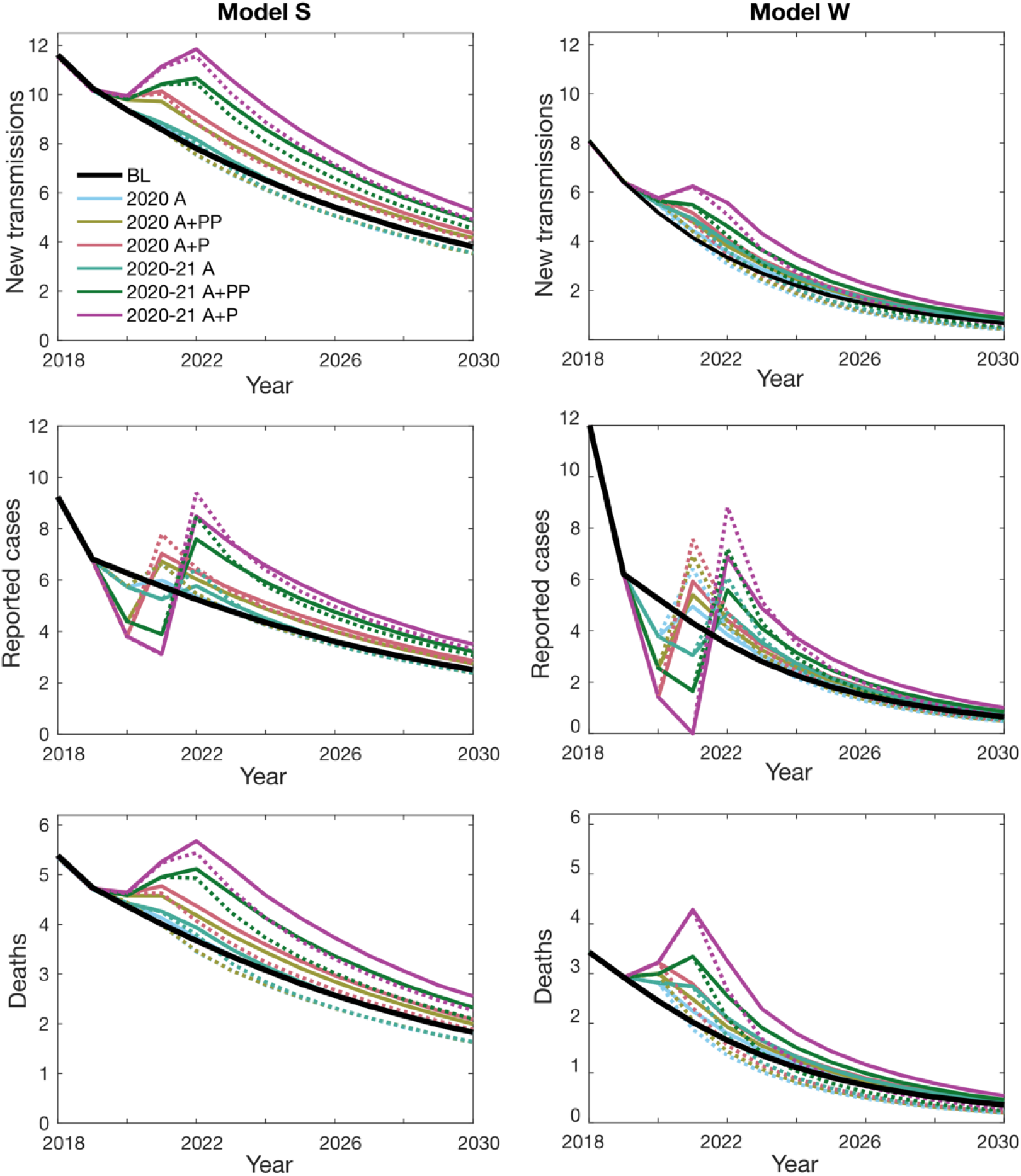
Predicted gHAT infections in the health zone Mosango based on different COVID-19 interruption scenarios. The graphs show the expected number of new transmissions, reported cases, and number of deaths caused by the disease (mean values), for Model S (left hand side) and Model W (right hand side). The baseline (BL) is shown as a black solid line. Individual interruption scenarios without mitigation are depicted by solid lines with different colours as indicated in the first panel. The corresponding mitigation scenarios are shown with dashed lines using comparable colours.

The impact of interruptions of screening programs was studied using two independently-developed stochastic models of gHAT infection, named Model S and Model W, developed independently and previously described in other modelling studies (Stone 2015, Rock 2015, Castano 2019, Crump 2020, Davis 2019). Both these models take into account different stages of the disease and transmission between vectors and humans who might be at low- or high-risk of exposure to vector bites. They also allow for various screening programmes, including passive surveillance (PS) and AS, through which infected people may be identified, and therefore treated (More details in SI and in (Castano 2020, Crump 2020). A table with the similarities and differences in the models and simulations can be found in the SI.

The parameters used in the stochastic models were taken from fitting of corresponding deterministic models to the human case screening data recorded for the years 2000-2016 in each health zone, except for the Model S calibration for Bagata, which used data for 2000-2018 (see (Crump 2020) and SI). The fits assumed a 3% annual increase of the human population according to the available estimates, and an increase in the passive detection rate over time - an improvement that is backed by both anecdotal evidence and previous modelling studies (Crump 2020 and Castano 2020 PLoS (data aggregation)). Annual rates of active screening were captured directly from the data for the period of 2000-2018. The stochastic simulations use 200 parameter sets of the posterior distributions of specific health zones, each model generates 200,000 stochastic (1000 from each of the 200 parameter sets) and the model outputs compared to historic data can be seen in the SI (Figures 2 and 4).

### Interruption Scenarios

For a no-interruption baseline, we assume AS and PS interventions continue indefinitely from 2019 with the same number of people screened annually, given by the mean value of the last five years (2014-2018). We then consider six potential interruption scenarios of gHAT activities due to COVID-19. We assume all interruptions start at the beginning of April 2020, but they may last until either the end of 2020 or 2021. The interruptions may disrupt either AS or both AS and PS. Whilst AS is assumed to be fully suspended within the interruption period, PS may be partially operating, going back to the detection capacity before modelled improvement; for Model W this was the PS level in 1998, and in Model S this was for 2000. Table 1 summarises these six interruption scenarios. The interventions are reinstated to the baseline values (mean AS and full PS) after the interruption period. Moreover, we study identical scenarios with mitigation, where AS is set to the maximum coverage observed in the data between 2000 and 2018, after interruption finishes. Simulating these 13 scenarios allows us to study possibilities of catching up to previously expected progress or even accelerating towards the 2030 goal.

**Table 1.** gHAT strategies and interruption scenarios (due to COVID-19) considered in this simulation study.

| Name | Interruption length | Active screening (AS) during interruption | Passive surveillance (PS) during interruption | Mitigation after interruption |
| --- | --- | --- | --- | --- |
| Baseline (BL) | None | Mean of 2014-2018 | Full (current detection rate is fitted) | NA |
| 2020 A | From April 2020 until end of 2020 | None | Full | No: Mean AS reinstated<br>Yes: Max AS begins |
| 2020 A + PP | From April 2020 until end of 2020 | None | Partial (levels in 2000 for Model S and 1998 for Model W) | No: Mean AS reinstated<br>Yes: Max AS begins |
| 2020 A + P | From April 2020 until end of 2020 | None | None | No: Mean AS reinstated<br>Yes: Max AS begins |
| 2020-21 A | From April 2020 until end of 2021 | None | Full | No: Mean AS reinstated<br>Yes: Max AS begins |
| 2020-21 A + PP | From April 2020 until end of 2021 | None | Partial | No: Mean AS reinstated<br>Yes: Max AS begins |
| 2020-21 A + P | From April 2020 until end of 2021 | None | None | No: Mean AS reinstated<br>Yes: Max AS begins |

## Results

We compare different scenarios by following the predicted infection dynamics over time after the interruptions are introduced in both health zones. In particular, our model outputs include the annual number of new human infections, reported cases corresponding to both passive surveillance and active screening, and the deaths caused by gHAT. We calculate these variables for all realisations and examine at the mean values of all 200,000 realisations. Figures 1 and 2 show the mean dynamics over time of all 13 scenarios as introduced in Table 1. In some cases the mean is substantially higher than the median and so we provide baseline projections in the SI showing both averages and also 95% prediction intervals for the two models and both health zones (see SI Figures 3 and 6). These wider prediction intervals explain how the mean new transmissions in Model S can be higher than Model W for Mosango (Figure 2), however there is a higher probability of meeting EOT during 2020-2025 for Model S (Figure 3).

**Fig 4.**
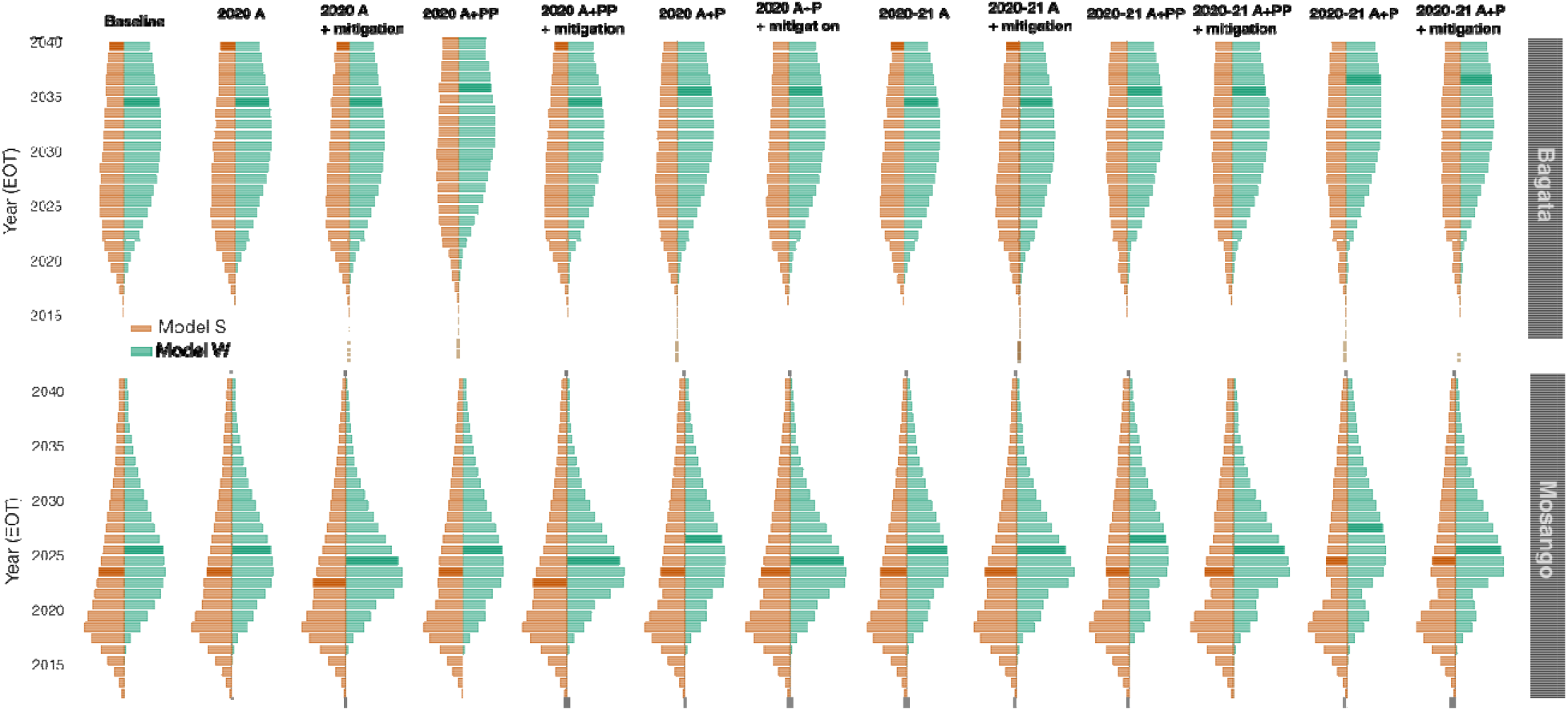
Distribution of the year in which EOT is predicted under different interruption scenarios. The violin plots compare the probability of EOT being achieved during each year and show Model W (green) and Model S (orange). Larger bars have higher probabilities of EOT being achieved in that year and the median elimination year is shown by a darker colour.. The first and second rows correspond to Bagata and Mosango respectively. N.B. Probabilities after 2040 were not computed, however several of the Model S simulations for Bagata have median EOT years after 2040 and the darker shaded median bars are not displayed in those cases.

**Fig 3.**
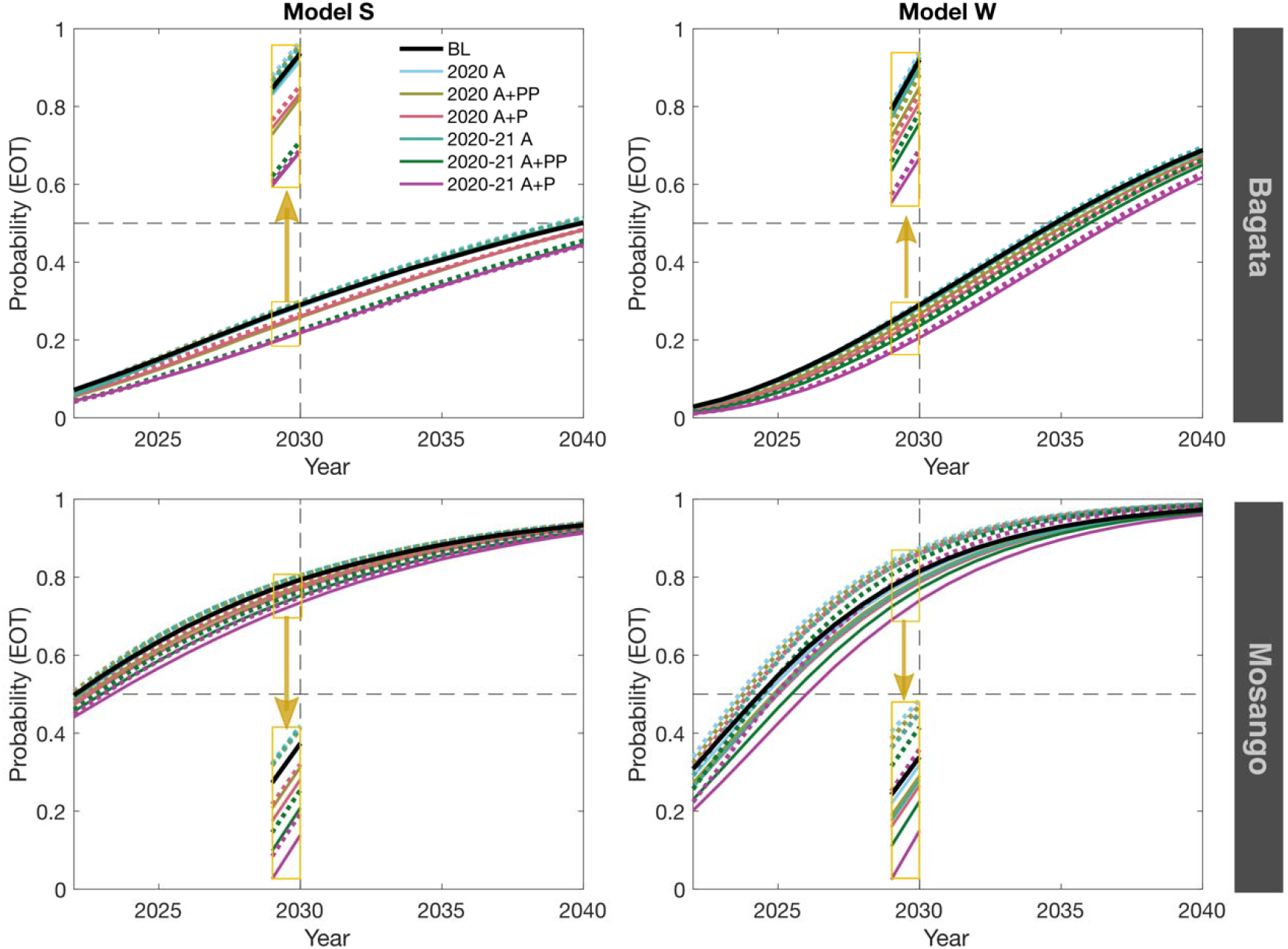
Cumulative probability of elimination of transmission by year. Predictions of both models showing the probability of achieving EOT by a given year are plotted for the period of 2022-2040 for the health zones Bagata and Mosango under different interruptions and the corresponding mitigation scenarios (maximum AS after the interruption) are shown by dashed lines. A zoomed version around 2030 is shown as inset (yellow boxes).

Similar trends can be recognised for the two health zones Bagata and Mosango despite their different prevalence and infection dynamics. The results from both models and both health zones predict a significant increase in the mean number of new infections following suspension of both AS and PS. On the other hand, the number of reported cases decreases during the interruption period, but then resurges in the following years, especially in mitigated scenarios where active screening is reinstated at the maximum historic coverage. More importantly, our models predict higher death rates during and after the interruptions. If PS continues, either at full or reduced capacity, our simulations indicate that infection is unlikely to increase during the interruption, however progress towards elimination of transmission could slow or stagnate during interruption. Putting these results together, the loss is more pronounced for longer interruptions (until the end of 2021) and, as expected, the worst response is observed when both active and passive activities are fully ceased. These results suggest retaining a minimum level of PS plays a significant role in controlling the transmission even if planned AS cannot go ahead. In the worst scenario, if all activities are suspended for two years, mitigations are not expected to help catch up with the baseline before 2030.

In addition to examining the expected infection and reporting dynamics, we also estimate the EOT probability by calculating the cumulative fraction of simulations reaching EOT over time for these health zones. In case of no interruptions, our models predict a fairly high probability (82% chance for Model W, and 77% for Model S) to achieve EOT in Mosango by 2030. Suspension of AS alone is not predicted to disturb that substantially (see Figure 3). This probability is clearly decreased, yet still above 50%, in the worst case scenario when all activities are suspended for two years even without mitigation. The median elimination year is delayed for almost three years in this scenario. Mitigated programs can facilitate elimination, and may even result in higher probabilities of achieving the EOT goal compared to the baseline scenario without interruption in the case that some PS was retained. In contrast to Mosango, reaching the 2030 EOT goal does not seem likely with the current programs in Bagata (only with 29% probability for both models), even without any pause in gHAT related activities. However, EOT by 2030 becomes more unlikely when severe interruptions are introduced. Our predictions for Bagata suggest the median elimination year between 2034-2035 which can be delayed to 2038-2039 in the worst scenario.

To visualise the predicted delays in elimination, we present violin plots representing the probability that EOT is predicted to occur during a specific year (probability density function of EOT) in our simulations (Figure 4), which clearly show how the distributions are shifted toward later years for severe scenarios.

Whilst the qualitative trends and even 2030 EOT probabilities for the two health zones are similar between models, we note that there are quantitative differences. In particular Model S has wider prediction intervals than Model W (see SI Figures 2 and 4) and this is reflected in both the higher means for new transmission in both health zones, even though medians are lower, and in the wider violin plot distributions.

## Conclusions and discussion

Our analysis of two independent models highlights the possible damage caused by the suspension of gHAT interventions due to COVID-19. In the most severe scenario we assume both AS and PS would be completely stopped, which is predicted to delay EOT by an average of 2-3 years if interruption continued until the end of 2021. However, the COVID-19 pandemic continues to evolve and it remains unclear when public health measures will be relaxed (or reinstated). In case of longer interruptions (not simulated here), we might expect more serious damage to the gains made to date by the elimination efforts.

The predictions made are specific to the two health zones of the DRC: Mosango and Bagata; other health zones could behave differently especially if current controls are playing a stronger role in reducing transmission. Our analysis predicts EOT may still be achieved in Mosango by 2030 thanks to the recent boost in AS coverage; however, in Bagata the elimination goal is unlikely without intensifying interventions, even without COVID-19 related interruptions. The heterogeneous nature of active screening in gHAT endemic areas (Franco, 2020) and the underlying focal nature of disease transmission (Franco, 2014) mean that results will be region-specific for the delay in EOT and the necessity of mitigation strategies should be evaluated on this basis. Additionally, in Mosango and Bagata there has been no wide-scale vector control, and we would expect qualitatively different impacts in areas where this has already been implemented. In the Ebola outbreak of 2014–2015, the interruption to active screening and passive surveillance is thought to have caused an increase in transmission in the affected areas, except in places where small insecticide impregnated targets could be maintained, indicating tsetse control is likely protective during medical interruptions if it remains in situ (Kagabadouno, 2018, Camara, 2017).

Moreover, our results suggest that retaining functioning passive surveillance, even partially, can help to avoid significant delays in EOT and to prevent substantial increases in mortality. Even with a functional health system, it is unclear how pandemic-induced changes in health-seeking behaviour, in addition to redirecting limited health resources, will impact levels of passive-case finding. Mitigation through increasing coverage of AS following interruption could also increase the probability of meeting the 2030 EOT goal. On the whole, our results suggest a milder impact of COVID-19 related delays on gHAT incidence and mortality than that suggested by similar studies on malaria (Sherrard-Smith 2020, Hogan 2020).

The use of stochastic events in both presented model formulations enable the direct computation of EOT prediction within health zones, by calculating the probability from multiple realisations of the infection process. However, we note that neither model considers the impact of an asymptomatic reservoir of human infection that is able to self cure or how an animal reservoir may be able to sustain transmission, as the extent of these factors remains unknown (Buscher, 2018, Capewell, 2019). Furthermore, local EOT for health zones will somewhat depend on EOT in neighboroughing regions, especially if there is substantial movement of people between locations. The modelling work here assumed that no importation of infection occurs, with health zones considered “epidemiological islands”. Previous modelling work indicates there may be only small rates of importations between villages in endemic regions within the Kwilu province (Davis, PloS NTD, Village scale), but nearby regions of continued transmission could pose a threat of re-introduction into health zones which successfully achieve EOT.

The use of two different models has enabled us to account for some structural and parameter uncertainty due to the way the models were constructed and fitted to observed data. A brief summary of the model differences is given in the SI (Table 5). Through fitting to data both models estimated the passive detection rates in 2000 and 2016 (see SI Tables 1 and 5), with both models achieving comparable values for 2016 -- those used for model projections -- in both health zones. There was a noticeable difference in inferred base passive detection values for Bagata between both models, leading to Model S estimating a ∼40-fold improvement between 2000 and 2016 for both stages, while Model W suggested that these improvements were more modest at around 4 times for stage 1 and 1.1 for stage 2. These structural and parameter differences illustrate why it is unsurprising that the two models provide different projections for the baseline and interruption scenarios, however despite this, they do reach consensus that the interruptions considered here would be unlikely to represent a large setback for the programme. Furthermore they provide similar quantitative estimates for the probability that each health zone will meet the EOT target by 2030 under baseline and suggest that Bagata in particular will need to have an intensified strategy to bring forward EOT and achieve the goal.

These results provide a rather optimistic perspective on COVID-19-related interruption on gHAT control. We stress that the cumulative impact of simultaneous programme interruptions for multiple diseases (NTDs and other infections) afflicting the same vulnerable populations could become an additional global health issue that deserves early attention and sustained control efforts.

## Supporting information

Model Description

Model S Fitting

## Data Availability

All the data used for this piece has been used in publish articles and the references and details are included in the SI.

## Declaration of author contributions

| Author's Initials | Study design | Study implementation | Analysis and interpretation of data | Major contribution to writing | Read and approved final version |
| --- | --- | --- | --- | --- | --- |
| MA |  | X | X | X | X |
| SC |  | X | X |  | X |
| CND |  |  | X | X | X |
| SP |  |  | X |  | X |
| EMM | X |  |  |  | X |
| SEFS |  |  | X |  | X |
| NC | X |  | X |  | X |
| MJK |  |  | X |  | X |
| KSR | X |  | X | X | X |

### Acknowledgements

The authors thank PNLTHA for original data collection, WHO for data access (in the framework of the WHO HAT Atlas \cite{Franco2018}). Calculations of model S were performed at sciCORE (http://scicore.unibas.ch/) scientific computing center at University of Basel.

