## Supplementary material for "Predicting the impact of COVID-19 interruptions on transmission of gambiense human African trypanosomiasis in two health zones of the Democratic Republic of Congo": Model Description

#### 1 Description of Model S

The Model S used in this study is a stochastic version of a variant of the ordinary differential equations (ODE) model presented in (1), which is based on the initial model published in (2). The model describes the transmission dynamics of *gambiense* human African trypanosomiasis (gHAT), and consists of a system of coupled ODEs, with compartments for tsetse, animal and human populations. These three different host types are modelled for two different settings corresponding to a low transmission area (e.g. the village, $L$) and a high transmission area (such as river banks or plantations, $H$) that enable accounting for heterogeneity in exposure to tsetse bites. The population size for tsetse, animal or humans in each setting $i$ ($i \in \{L, H\}$) is assumed to be stable by allowing the associated birth terms to compensate deaths in all the compartments. Tsetse and animal populations always stay within their setting (for example, tsetse in low transmission settings always remain in the low transmission setting and animals in high transmission settings always remain in the high transmission setting). Whilst humans in low transmission settings always remain in low transmission setting, humans in the high transmission setting move back and forth between the high and low transmission settings spending a fixed amount of time in each one (to model, for example, the movement of high risk individuals between villages and plantations) — as shown in Figure 1.

Five compartments describe humans in any of the two settings: susceptible ($S_{hi}$); exposed or incubating ($E_{hi}$); infected with the first stage of the disease ($I_{h1i}$); infected with the second stage of the disease, where trypanosomes have reached the cerebro-spinal fluid ($I_{h2i}$); and removed ($T_{hi}$). The total human population in setting $i$ is $N_{hi} = S_{hi} + E_{hi} + I_{h1i} + I_{h2i} + T_{hi}$.

Tsetse populations are divided into susceptible ($S_{vi}$); teneral ($U_{vi}$); exposed ($E_{vi}$); and infected ($I_{vi}$), so that the vector population is $N_{vi} = S_{vi} + U_{vi} + E_{vi} + I_{vi}$. Infected humans in both infected stages can transmit the parasite to tsetse flies. Animals do not contribute to transmission, thus animal populations are modelled as constant parameters, $N_{ai}$, and only form a sink for tsetse bite. A schematic of the model is shown in Figure 1.

The deterministic ODE version of Model S was fitted to health-zone-level data using an adaptive Metropolis-Hastings Markov chain Monte Carlo (MCMC) algorithm (see SI Model S - model fit).

Stochastic simulations in this analysis used 200 posterior parameter sets sampled from a total of 10,000 samples. To achieve reasonable statistics, we performed 1,000 realisations of the stochastic model for each parameter set.

In this study, the model dynamics is described by random events captured by an adaptive tau-leap algorithm (3), implemented using the 'adaptivetau' R package (4) with the default setting. This adaptive scheme defines the time increment of the simulations in an adaptive manner depending on the state of the system at each time step. The adaptive tau-leaping algorithm dynamically switches between three methods for simulating events: either an explicit or implicit tau-leaping (3), in which the number of events taking place in a time interval $\tau$ is chosen randomly from a Poisson distribution with the mean given by the event rate defining transition, multiplied by time step $\tau$; or an exact method, that is Gillespie's direct method (5), when the leap condition is violated, thus avoiding the issue of negative populations that can happen with tau-leap methods.

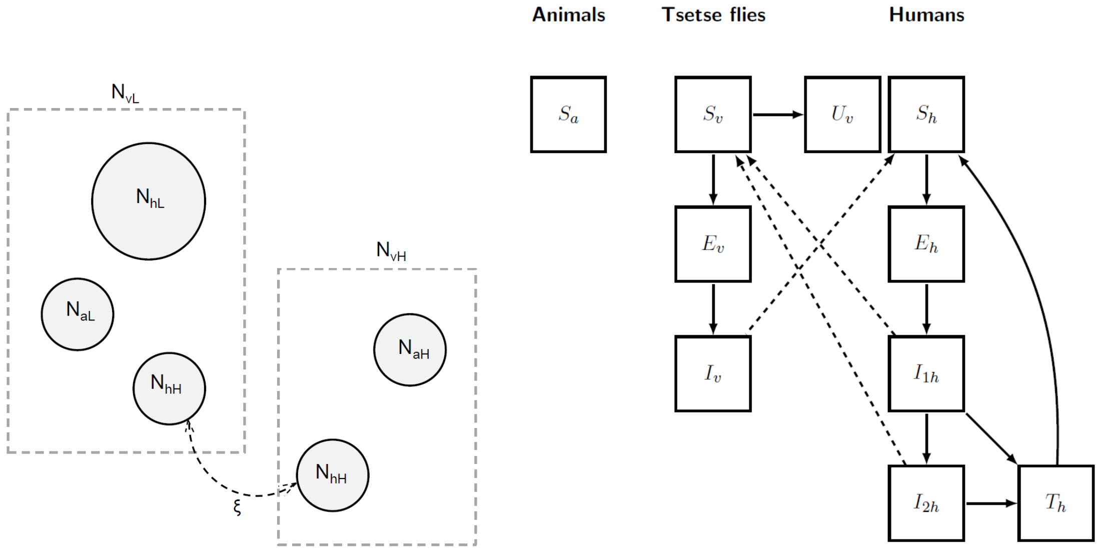


Figure 1: Schematic of Model S. Left: model population structure. Human populations are composed by a stationary population ($N_{hL}$) that remains in low exposure habitats (e.g., a village), and a smaller population ($N_{hH}$) which commute and spend a proportion $\xi$ of their time in a potentially high exposure setting (e.g., a plantation). Each habitat also contains tsetse ($N_{vL}$ and $N_{vH}$) and non-human vertebrate animal populations ($N_{aL}$ and $N_{aH}$). Right: schematic of infection dynamics, subscripts $i \in \{L, H\}$ were removed for easy reading. Compartmental diagram highlights the transmissions between states of infection of the tsetse and human populations, with solid lines indicating transition between compartments, and dashed lines representing transmission rates. Animals cannot transmit infection, thus acting as a sink for tsetse bite. Note that in the low-risk transmission setting, both human populations are exposed to tsetse bites. Figure adapted from (2).

#### 1.1 Screening

Active screening is modelled via a constant annual passive detection rate $r_{as}$ that removes infected people only from the low risk setting. As in previous works, we followed (6) to relate a proportion, $d$, of humans effectively screened in a give year and the annual removal rate $r_{as}$ as $d = 1 - e^{-r_{as}}$, leading to $r_{as} = -\ln(1-d)$. For the model fitting, screening levels were informed from data, and estimates for the health zone population in 2015 were taken from (7), and projected backwards and forward in time assuming a 3% annual growth rate. For model projections, the mean number of people screened from the last 5 years of available data (2014-2018) was used to define $d$, with an ongoing 3% growth rate in the total population (leading to a continuing decrease in the proportion of the population screened).

Passive detection is represented by a continuous stage-specific detection rate, $r1$ and $r2$ for stage 1

| | Mosango [2.5, 25, 50, 75 and 97.5% CIs] | Bagata [2.5, 25, 50, 75 and 97.5% CIs] |
|---|---|---|
| Stage 1 rate (2000) | $10^{-3} \times [0.193, 0.253, 0.288, 0.324, 0.389]$ | $10^{-5} \times [1.739, 2.325, 2.653, 3.017, 4.003]$ |
| Stage 1 rate (2016) | $10^{-3} \times [0.582, 0.721, 0.807, 0.909, 1.11]$ | $10^{-3} \times [0.585, 0.854, 1.037, 1.233, 1.571]$ |
| Stage 2 rate (2000) | $10^{-3} \times [0.461, 0.666, 0.825, 1.02, 1.44]$ | $10^{-5} \times [3.691, 5.054, 5.886, 6.821, 9.072]$ |
| Stage 2 rate (2016) | $10^{-3} \times [1.167, 1.853, 2.360, 2.991, 4.708]$ | $10^{-3} \times [1.239, 1.922, 2.509, 3.002, 3.843]$ |

Table 1: Model S passive detection rates (in days$^{-1}$) over time

and stage 2 respectively, and removes infected people from both low- and high-risk settings. Relying on previous work (8), improvement to passive detection was assumed for the data period. We modelled improvement for the number of years $y > 0$ after 2000 as a logistic function:

$$r1(y) = 365 \times r1_{\text{const}} + \frac{\Delta r_1}{1 + \exp(-\alpha_{\text{pd}}(y - x_0 + 1))},$$

$$r2(y) = 365 \times r1_{\text{const}} \times c2 + \frac{\Delta r_1 + \Delta r_2}{1 + \exp(-\alpha_{\text{pd}}(y - x_0 + 1))},$$

where $r1_{\text{const}}$ and $c_2 * r1_{\text{const}}$ are the constant daily passive detection rates in stage 1 and stage 2 respectively for any time before 2000, and $\Delta r_1$, $\Delta r_2$, $\alpha_{\text{pd}}$ and $x_0$ are parameters defining the profile of the logistic curve. All parameters in the expression above are fitted to the health zone level data. A summary of daily rates at the beginning and end of the data period used for fitting is given in Table 1. For model projections in the non-interruption scenario, we assumed passive detection rates $r1$ and $r2$ continue at the highest level from 2000-2016.

Our model assumes that before 2000 only passive detection was ongoing, at constant rates, and that active screening activities started in 2000, the initial year for which there is available data on active screening.

#### 1.2 Estimating model parameters

Descriptions and values of all fixed parameters used in the model are given in Table 2. The remaining parameters were sampled from posterior distributions (200 samples out of 10,000 samples) by fitting the model to health zone case data. Further details in model S calibration are presented separately in the supplementary information. Description of fitted parameters with their posterior median and $95\%$CI are provided in Table 3.

Table 2: **Model parameterisation (fixed parameters).** Notation, a brief description, and the used values of fixed parameters in Model S.

| Notation | Description | Value |
|---|---|---|
| $\alpha$ | Rate at which tsetse become non-teneral (i.e. cannot get infectious) | 73 year$^{-1}$ |
| $A/H_1$ | Density of animals relative to humans in area $L$ | 1.35 |
| $A/H_2$ | Density of animals relative to humans in area $H$ | 1.5 |
| $b$ | Proportion of infective bites leading to infection in humans and animals | 0.433 |
| $c_h$ | Proportion of bites on an infective human that lead to a mature infection in flies | 0.065 |
| $c_{ai}$ | Proportion of bites on an infective animal of type $i$ that lead to a mature infection in flies | 0 |
| $\delta$ | Rate at which treated humans return to the susceptible class | 2.19 year$^{-1}$ |
| $\delta_a$ | Rate of loss of immunity in animal hosts | 0.73 year$^{-1}$ |
| $\eta$ | Rate at which hosts move from the incubating stage | 31.025 year$^{-1}$ |
| $f$ | Inverse of duration of feeding cycle; or biting rate | 121.545 year$^{-1}$ |
| $\gamma$ | Rate of progression to stage 2 in humans | 0.6939 year$^{-1}$ |
| $\gamma_{aL}$ | Rate of progression to the immune class in animal hosts of type $L$ | 1 year$^{-1}$ |
| $\gamma_{aH}$ | Rate of progression to the immune class in animal hosts of type $H$ | 0.6935 year$^{-1}$ |
| $\mu$ | Death rate of humans due to natural causes | 0.01666 year$^{-1}$ |
| $\mu_{ai}$ | Death rate of animal host of type *i* | 0.584/0.6935 year$^{-1}$ |
| $\mu_\gamma$ | Disease-induced death rate or rate of leaving the recovered state for humans | 1.4484 Year$^{-1}$ |
| $\mu_t$ | Death rate of humans due to treatment | 0 year$^{-1}$ |
| $\mu_v$ | Death rate of tsetse | 10.95 year$^{-1}$ |
| $\nu$ | Inverse of the extrinsic incubation period | 12.41 year$^{-1}$ |
| $\sigma$ | Biting preference for humans | 0.326 |
| $\sigma_{ai}$ | Biting preference for animal in the setting $i$ | 0.8/0.396 |
| $\xi$ | Proportion of time spent i the high risk region by commuters | 0.698 |
| sensitivity | Diagnostics sensitivity (active screening) | 0.91 |

Table 3: **Model parameterisation (posteriors of fitted parameters).** Notation, a brief description, and representative percentiles of the posterior distributions for fitted parameters.

| Notation | Description | Posterior (median [95% CI]) Bagata | Mosango |
|---|---|---|---|
| $\kappa$ | Ratio of humans in the high- to low-exposure environment | 7.03 $\times 10^{-4}$ [ 1.27 , 20.5]$\times 10^{-4}$ | 4.52 $\times 10^{-3}$ [3.41, 5.98]$\times 10^{-3}$ |
| log(VHL) | Log ratio of vectors to humans in low-exposure environment | 1.359 [1.312, 1.386] | 1.180 [1.159, 1.207] |
| log($c_1$) | Log ratio of VHH to VHL | 8.58 $\times 10^{-2}$ [0.43, 89.29]$\times 10^{-2}$ | 2.44$\times 10^{-2}$ [0.099, 10.26]$\times 10^{-2}$ |
| logit(spec) | Diagnostic specificity (active screening) | 8.815 [8.382, 9.183] | 6.571 [6.331, 6.807] |
| $r1_{\text{const}}$ | Daily passive detection rate for stage 1 (pre-2000) | 3.33$\times 10^{-6}$ [0.28, 12.6]$\times 10^{-6}$ | 1.33$\times 10^{-5}$ [0.042, 5.35]$\times 10^{-5}$ |
| log($c_2$) | Log ratio of passive detection for stage 2 to stage 1 (pre-2000) | 0.331 [0.015, 0.953] | 0.466 [.0134, 2.163] |
| $\Delta r_1$ | Amount passive detection in stage 1 improves | 0.519 [0.223, 0.106] | 0.289 [0.207, 0.404] |
| $\Delta r_2$ | Amount passive detection in stage 2 improves (in addition to improvement of stage 1) | 0.704 [0.146, 1.766] | 0.554 [0.0838, 1.484] |
| $x_0$ | Turning point (years since 1999) for logistic improvement in passive detection | 13.75 [9.63, 17.68] | 2.63 [1.50, 4.33] |
| $\alpha_{\text{pd}}$ | Steepness in logistic improvement of passive detection | 0.327 [0.280, 0.390] | 0.405 [0.260, 0.623] |
| $\kappa_{\text{as}}$ | Overdispersion parameter (active screening) | 2.01 [1.45, 2.93] | 35.98 [20.88, 59.29] |
| $\kappa_{\text{pd}}$ | Overdispersion parameter (passive detection) | 48.96 [29.81, 76.06] | 66.06 [42.01, 97.58] |

#### 1.3 Model S stochastic simulation results

Figure 2 shows 200,000 stochastic realisations of the model (2000 parameter sets and 1000 realisations per set) against historic data for the two health zones considered in the present study. Projections for 2018-2040 are shown in Figure 6.

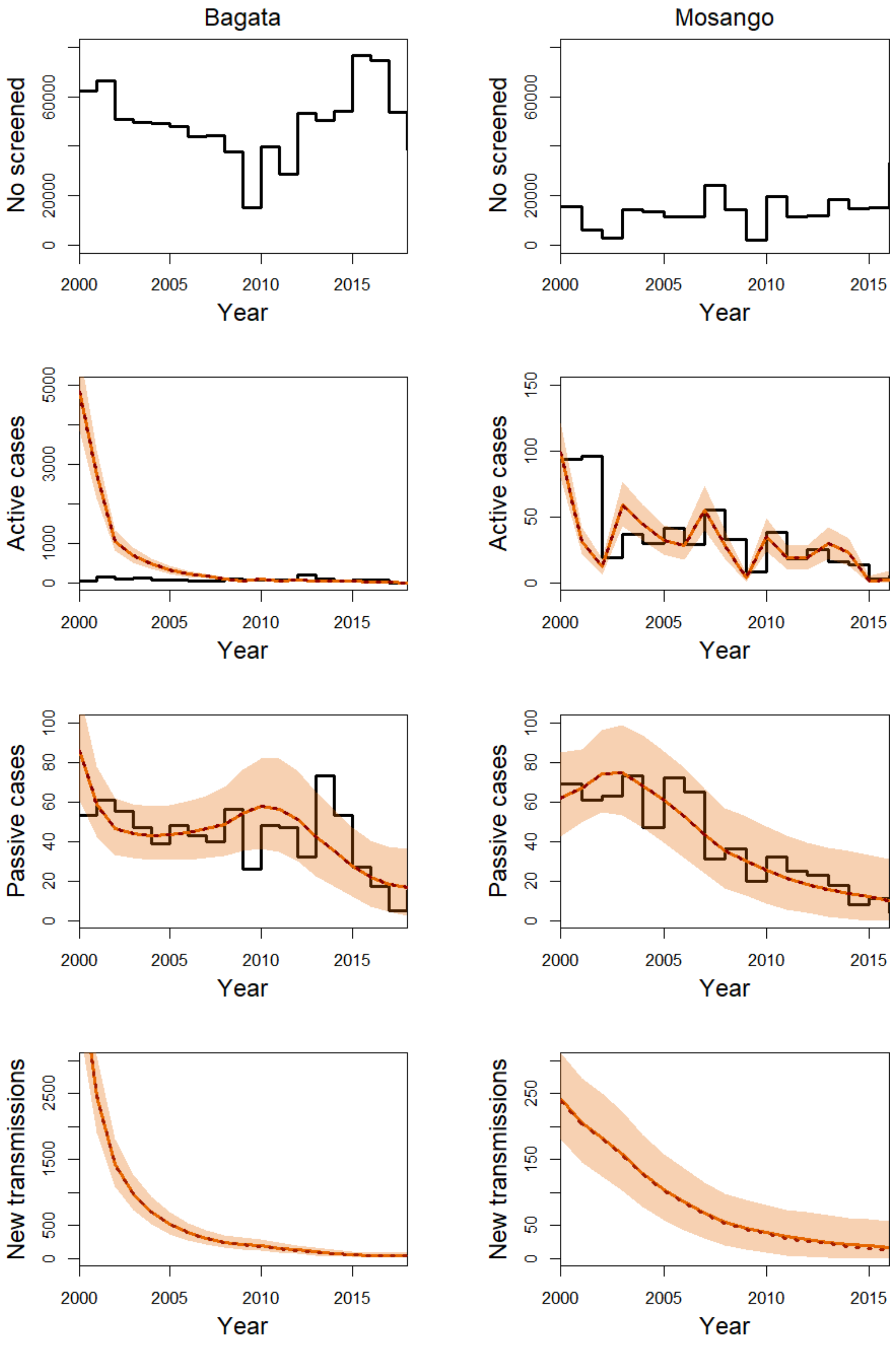


Figure 2: Time series plots of Model S against historic case data for health zones Bagata (2000–2018) and Mosango (2000–2016). The top row shows historic active screening coverage, the second row shows active case detections, the third shows passive case detections, and the bottom give the inferred transmission dynamics. The data is shown as black lines, with coloured solid and dotted lines showing the model outputs (mean = solid, median = dotted) and 95% credible intervals are the shaded region.

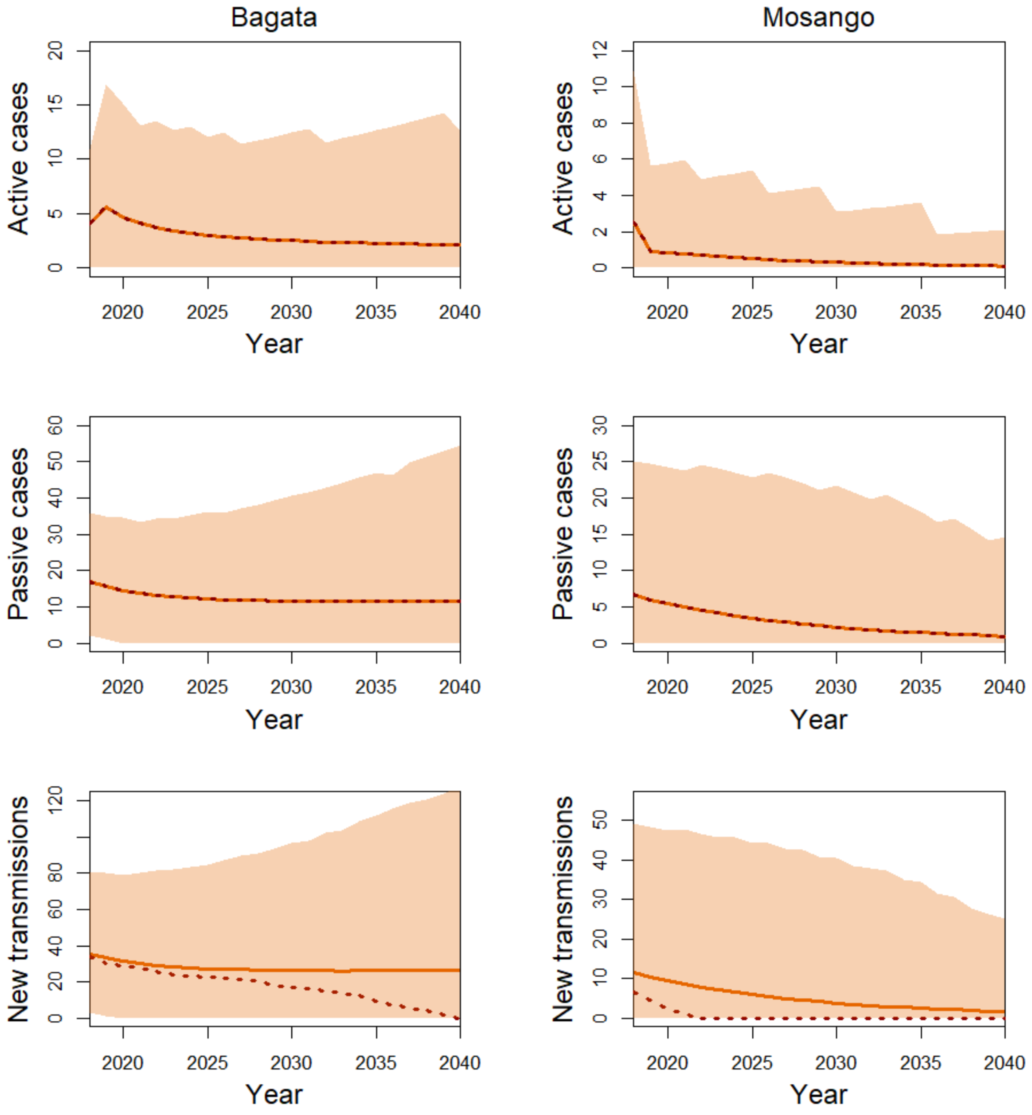


Figure 3: Time series plots of Model S projections for Bagata and Mosango health zones(2018–2040), showing active case detections (first row), passive case detections (second row), and transmission dynamics (third row). Mean (lines) and median (dots) as well as 95% prediction intervals (shaded area) are shown.

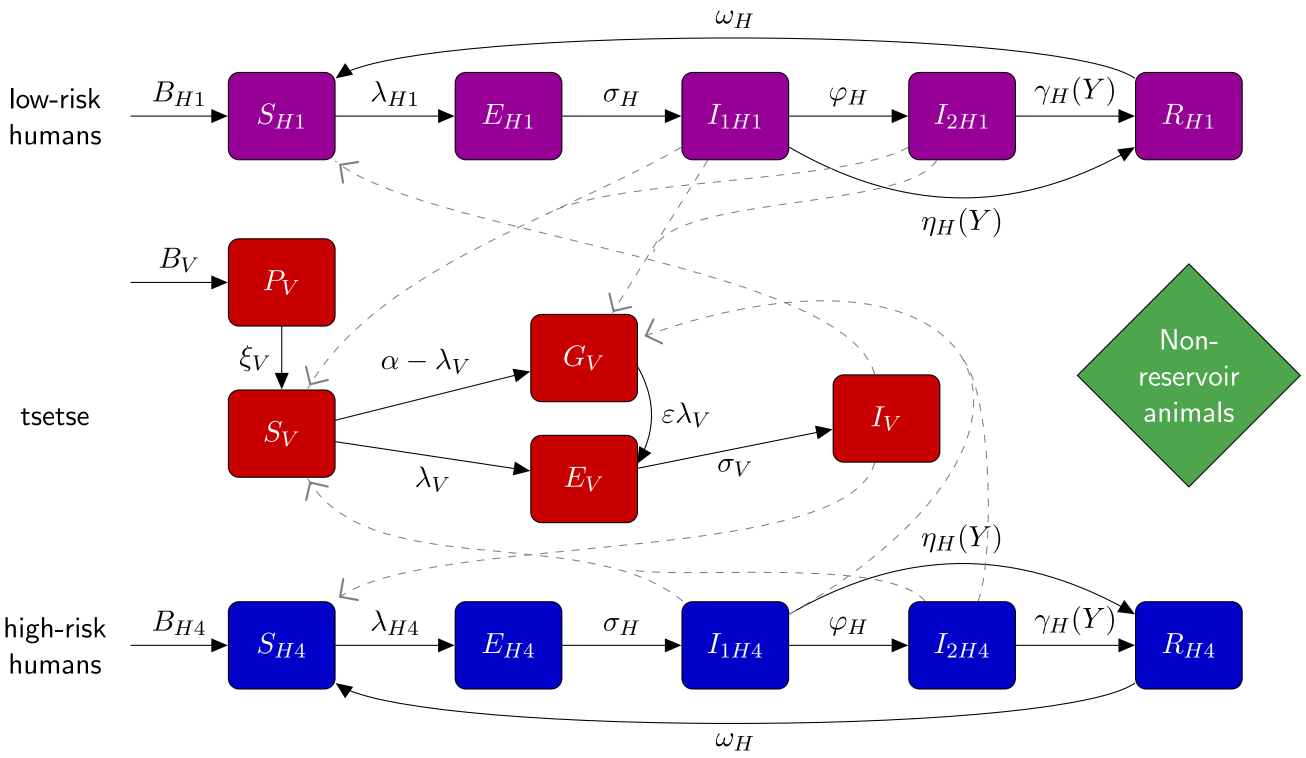


Figure 4: **Schematic of the model to describe gHAT infection dynamics.** This multi-host model of HAT takes into account high- and low-risk groups of humans and their interactions with tsetse vectors. Each group consists of different compartments: Susceptible humans $S_{Hi}$ can become exposed on a bite of an infectious tsetse. Exposed people $E_{Hi}$ progress to become the stage $1$ infected people and eventually stage $2$ (if not detected in screening), and once treated they recover by hospitalization $R_{Hi}$. Active screening can accelerate treatment rate of infected people. Here we assume high-risk group does not participate in active screening. By biting an infectious person, tsetse can become exposed and subsequently infectious, $E_V$ and $I_V$. $G_V$ represents the tsetse population not exposed to *Trypanosoma brucei gambiense* in the first blood-meal and are therefore less susceptible in the following meals. Rates are shown by Greek letters associated with arrows. Animal reservoir is not considered. This figure is taken from (9) and adapted from the original model schematic (10).

#### 2 Description of Model W

Our model to describe gHAT dynamics is based on the ODE model presented in (10; 9) and the extended stochastic version in (11; 1). Figure 4 shows a schematic description of gHAT dynamics in this model that takes into account different compartments of humans and tsetse. Humans can be exposed and subsequently infected by a bite of an infectious tsetse. They progress through stage 1 and stage 2 of the infection with specific rates. On the other side, tsetse vectors can become exposed and subsequently infectious if they bite an infectious human. Infected people may be detected by passive and active screening (see below for more details), followed by hospitalisation and recovery. Here, we consider a version of the model where humans are partitioned into two sub-groups of (i) low-risk and participating in the active screening, and (ii) high-risk and non-participating in active screening. We assume there are no animal reservoirs although animals receive some proportion of tsetse bites.

In this stochastic model, individual humans are assigned to different compartments associated with infection/disease status and can transition between them. We describe system dynamics by random events captured by a tau-leap approximation. Table 4 explains different events and the corresponding rates that lead to one person transitioning from one compartment to another one. Within this framework, the number of events happening in a time interval $\tau$ is chosen randomly from a Poisson distribution with the mean equal to the event rate multiplied by $\tau$. We use time interval of a day in the tau-leaping algorithm that is shown to be a sensible choice for gHAT dynamics (11).

Table 4: Model W stochastic formulation (human dynamics)

| **Event** | **Transition** | **Rate** |
|---|---|---|
| Recovery from hospitalisation | $s_{Hi} \to s_{Hi} + 1, r_{Hi} \to r_{Hi} - 1$ | $\omega_H r_{Hi}$ |
| Natural death of hospitalised | $s_{Hi} \to s_{Hi} + 1, r_{Hi} \to r_{Hi} - 1$ | $\mu_H r_{Hi}$ |
| Exposure of susceptibles | $s_{Hi} \to s_{Hi} - 1, e_{Hi} \to e_{Hi} + 1$ | $f_{Hi} \alpha m_{eff} s_{Hi} I_V / N_{Hi}$ |
| Progression to Stage 1 infection | $e_{Hi} \to e_{Hi} - 1, i_{1Hi} \to i_{1Hi} + 1$ | $\sigma_H e_{Hi}$ |
| Natural death of exposed | $e_{Hi} \to e_{Hi} - 1, s_{Hi} \to s_{Hi} + 1$ | $\mu_H e_{Hi}$ |
| Progression to Stage 2 infection | $i_{1Hi} \to i_{1Hi} - 1, i_{2Hi} \to i_{2Hi} + 1$ | $\varphi_H i_{1Hi}$ |
| Natural death of Stage 1 infection | $i_{1Hi} \to i_{1Hi} - 1, s_{Hi} \to s_{Hi} + 1$ | $\mu_H i_{1Hi}$ |
| Treatment or death from Stage 2 infection | $s_{Hi} \to s_{Hi} + 1, i_{2Hi} \to i_{1Hi} - 1$ | $\gamma_H(Y) i_{2Hi}$ |
| Natural death of Stage 2 infection | $i_{2Hi} \to i_{2Hi} - 1, s_{Hi} \to s_{Hi} + 1$ | $\mu_H i_{2Hi}$ |

To avoid the need to disaggregate the parameter $m_{eff}$ - which was used to non-dimensionalise the previous ODE system - into its component parts ($p_H$ and $N_V/N_H$), we keep vector dynamics the same as the original model, described by a set of ODEs:

$$\begin{aligned}
\frac{dS_V}{dt} &= \mu_V N_H - \alpha S_V - \mu_V S_V \\
\frac{dE_{1V}}{dt} &= \alpha p_V (f_{H1} \frac{I_{1H1} + I_{2H1}}{N_{H1}} + f_{H2} \frac{I_{1H2} + I_{2H2}}{N_{H2}})(S_V + \varepsilon G_V) - (3\sigma_V + \mu_V) E_{1V} \\
\frac{dE_{2V}}{dt} &= 3\sigma_V E_{1V} - (3\sigma_V + \mu_V) E_{2V} \\
\frac{dE_{3V}}{dt} &= 3\sigma_V E_{2V} - (3\sigma_V + \mu_V) E_{3V} \\
\frac{dI_V}{dt} &= 3\sigma_V E_{3V} - \mu_V I_V \\
\frac{dG_V}{dt} &= \alpha \left(1 - p_V \left(f_{H1} \frac{I_{1H1} + I_{2H1}}{N_{H1}} + f_{H2} \frac{I_{1H2} + I_{2H2}}{N_{H2}}\right)\right) S_V \\
&- \alpha p_v \left(f_{H1} \frac{I_{1H1} + I_{2H1}}{N_{H1}} + f_{H2} \frac{I_{1H2} + I_{2H2}}{N_{H2}}\right) \varepsilon G_V - \mu_V G_V.
\end{aligned} \tag{2.1}$$

This is a legitimate assumption due to the high population of vectors and their short life cycle compared to humans. We solve these equations with the help of Runge-Kutta methods.

#### 2.1 Screening

This model accounts for the possibility of detecting of infected humans through passive and active screening. Passive screening describes potential visits of people to fixed medical centers for testing. In the stochastic version, we identify the number of people participating in passive screening by a tau-leaping approximation with rates proportional to $\eta_H$ and $\gamma_H$ corresponding to the first and second stages of the disease (Table 4).

Before 1998 (pre-active screening) it was assumed that passive detection was less effective than after activities began, and only so identified stage 2 individuals at a rate $\gamma_{H0}$, which is smaller than the stage 2 passive detection rate from 1998 onwards, $\gamma_H$. We assumed no stage 1 detections pre 1998, i.e. $\eta_H = 0$.

| | Mosango [2.5, 25, 50, 75 and 97.5% CIs] | Bagata [2.5, 25, 50, 75 and 97.5% CIs] |
|---|---|---|
| Stage 1 rate (2000) | $10^{-3}\times$[0.031, 0.076, 0.11, 0.15, 0.27] | $10^{-3}\times$[0.12, 0.18, 0.23, 0.3, 0.48] |
| Stage 1 rate (2016) | $10^{-3}\times$[0.08, 0.16, 0.22, 0.29, 0.46] | $10^{-3}\times$[0.5, 0.7, 1.0, 1.3, 2.0] |
| Stage 2 rate (2000) | $10^{-3}\times$[1.1, 1.9, 2.4, 3.2, 6.3] | $10^{-3}\times$[0.6, 1.0, 1.4 1.9, 3.5] |
| Stage 2 rate (2016) | $10^{-3}\times$[1.4, 2.4, 3.2, 4.4, 9.2] | $10^{-3}\times$[0.6, 1.0, 1.6, 2.1, 3.8] |

Table 5: Model W passive detection rates over time

Following previous modelling work using gHAT data from former Bandundu province, there is a strong signal from epidemiological staging data that passive screening has improved during the time period from 2000–2016 (8; 9). To capture the improvement of stage 1 to stage 2 passive detection, the model utilises the following formula:

$$\begin{aligned}
\eta_H(Y) &= \eta_H^{\text{post}}\left[1+\frac{\eta_H\text{amp}}{1+\exp\left(-d_{\text{steep}}(Y-d_{\text{change}})\right)}\right], \\
\gamma_H(Y) &= \gamma_H^{\text{post}}\left[1+\frac{\gamma_H\text{amp}}{1+\exp\left(-d_{\text{steep}}(Y-d_{\text{change}})\right)}\right],
\end{aligned}$$

where $Y$ is the year and $\eta_H(Y)$ is the annual stage 1 passive detection rate. Parameters dictating the amplitude, steepness and switching year can be found in Tables 6 and 7. The free parameters have been estimated through fitting to the health zone level data for Bagata and Mosango. A summary of rates at the beginning and end of the data period used for fitting is given in Table 5.

In our current approach, active screening is considered to be a random procedure to detect infected humans within the people participating voluntarily. The number of people picked up from each compartment is given by a random number chosen from a binomial distribution. Similar to the previous models, we allow for the imperfect nature of the tests by considering sensitivity of tests to detect true cases and specificity to observe false positive cases. Specificity is set to one after 2015 due to improvement in confirmatory quality control (9).

Using a similar approach to the previous ODE models, we consider the same level of screening as reported between 2000–2016. It is assumed, as in much of the previous published studies using this model, that active screening began in 1998 and achieved the same number of people screened as in 2000 (the first year of data). In 2017 and 2018 we used the screening values from the WHO HAT Atlas (even though data were not used for the fitting) and after 2018, we use the average percentage of screened people between 2014–2018 for different strategies (and maximum for mitigation scenarios).

#### 2.2 Estimating model parameters

As in previous versions of the deterministic model (10; 9), some parameters with estimates available in the literature were assigned fixed values. Fixed values are given in Table 6. The other parameter values were taken from posterior distributions by fitting the model to data. In this approach, the deterministic model was fitted to health-zone-level data for different health zones using an adaptive Metropolis-Hastings MCMC algorithm (9). In the present analysis, we used 200 parameter sets from the posterior distributions for two different health zones of Bagata and Mosango.

We use the stochastic version of the described gHAT model to study the extinction time (11; 1). In this stochastic picture, individual humans are assigned to different compartments associated with infection/disease status and can transfer between them. We describe system dynamics by random events captured by a tau-leap approximation. Table 4 explains different events and the corresponding

Table 6: **Model parameterisation (fixed parameters).** Notation, a brief description, and the used values of fixed parameters in model W.

| Notation | Description | Value |
|---|---|---|
| $N_H$ | Total human population size (in 2015) | M:121,433 B:165,990 (7) |
| $B_H$ | Total human birth rate | $= \mu_H N_H$ |
| $\mu_H$ | Natural human mortality rate | $5.4795\times10^{-5}$ days$^{-1}$ (12) |
| $\sigma_H$ | Human incubation rate | 0.0833 days$^{-1}$ (13) |
| $\varphi_H$ | Stage 1 to 2 progression rate | 0.0019 days$^{-1}$ (14; 15) |
| $\omega_H$ | Recovery rate/waning-immunity rate | 0.006 days$^{-1}$ (16) |
| Sens | Active screening diagnostic sensitivity | 0.91 (17) |
| $B_V$ | Tsetse birth rate | 0.0505$^*$ days$^{-1}$ (18) |
| $\xi_V$ | Pupal death rate | 0.037 days $^{-1}$ |
| $K$ | Pupal carrying capacity | $= 111.09 N_H^{\dagger}$ (18) |
| $\mathbb{P}$(pupating) | Probability of pupating | 0.75 |
| $\mu_V$ | Tsetse mortality rate | 0.03 days$^{-1}$ (13) |
| $\sigma_V$ | Tsetse incubation rate | 0.034 days$^{-1}$ (19; 20) |
| $\alpha$ | Tsetse bite rate | 0.333 days$^{-1}$ (21) |
| $p_V$ | Probability of tsetse infection per single infective bite | 0.065 (13) |
| $\varepsilon$ | Reduced non-teneral susceptibility factor | 0.05 (10) |
| $f_H$ | Proportion of blood-meals on humans | 0.09 (22) |

rates that lead to one person transitioning from one compartment to another one. Within this framework, the number of events happening in a time interval $\tau$ is chosen randomly from a Poisson distribution with the mean equal to the event rate multiplied by $\tau$. The same procedure is used to identify the number of people participating in passive or active screenings. We use time interval of a day in the tau-leaping algorithm that is shown to be a sensible choice for gHAT dynamics (11).

#### 2.3 Model W stochastic simulation results

Figure 5 shows 200,000 stochastic realisations of the model (2000 parameter sets and 1000 realisations per set) against historic data for the two health zones considered in the present study.

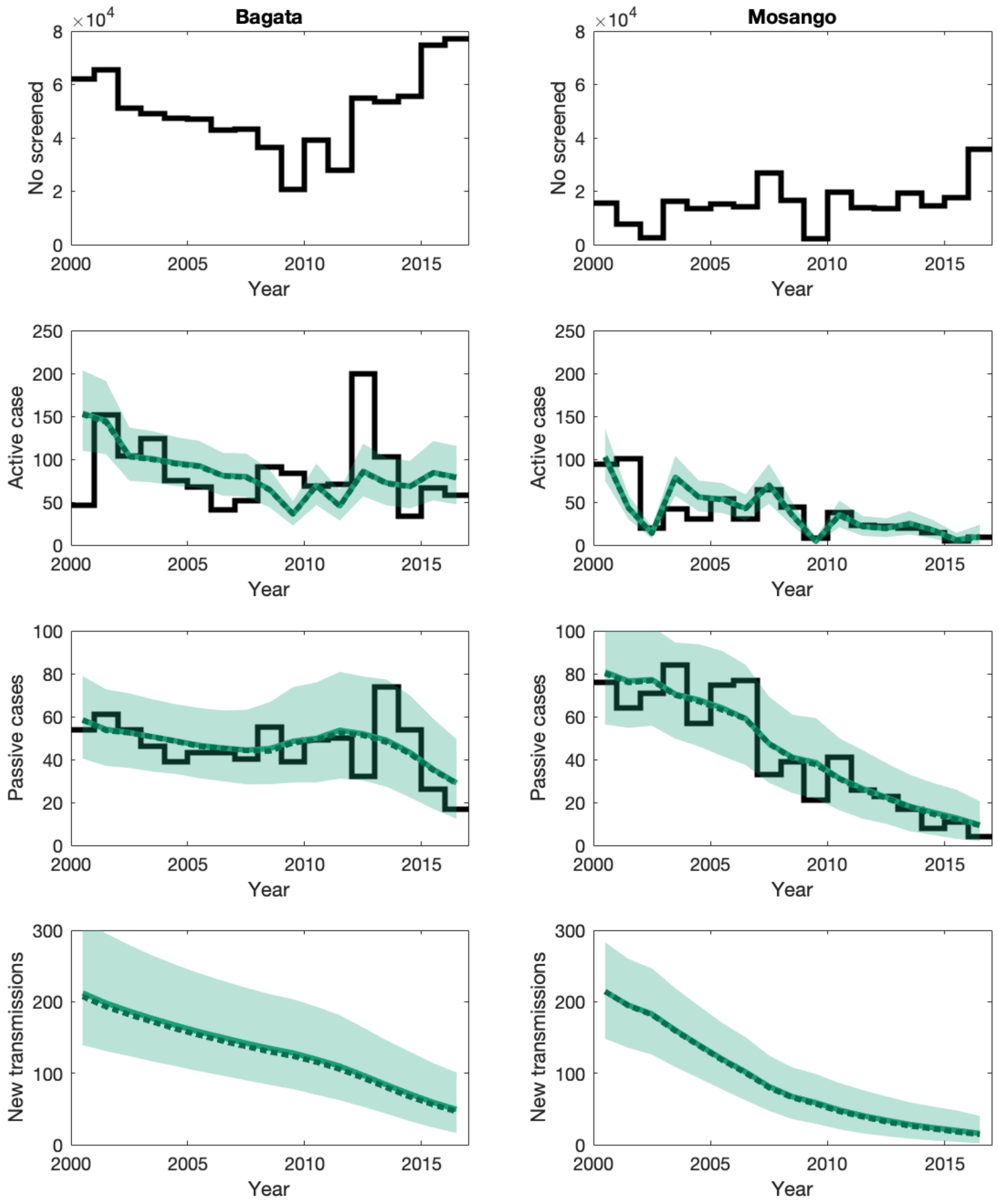


Figure 5: Time series plots of Model W against historic case data (2000–2016) for health zones Bagata and Mosango. The top row shows historic active screening coverage, the second row shows active case detections, the third shows passive case detections, and the bottom give the inferred transmission dynamics. The data is shown as black lines, with coloured solid and dotted lines showing the model outputs (mean = solid, median = dotted) and 95% credible intervals are the shaded region.

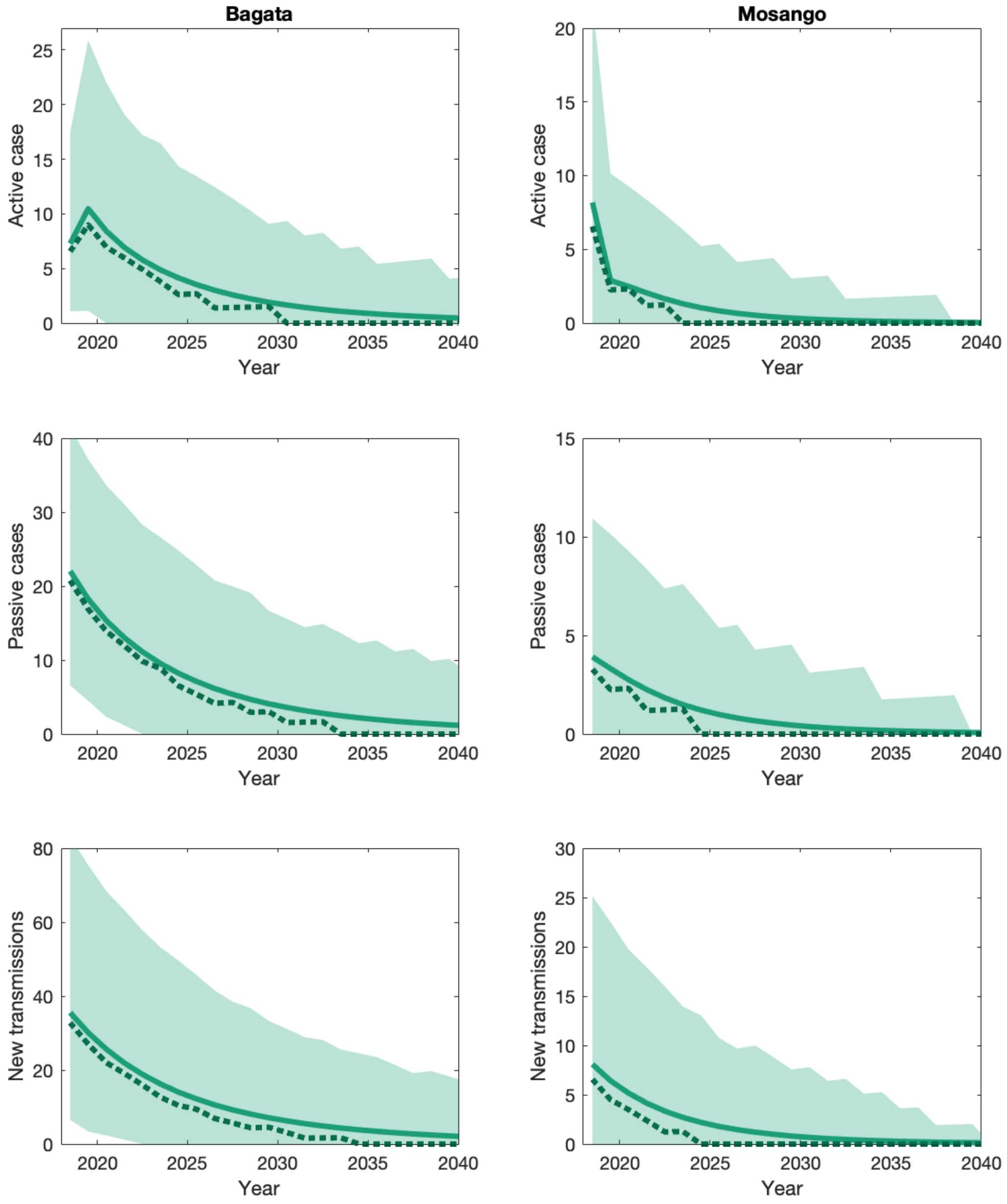


Figure 6: Time series plots of Model W projections for Bagata and Mosango health zones (2018–2040), showing active case detections (first row), passive case detections (second row), and transmission dynamics (third row). Mean (lines) and median (dots) as well as 95% prediction intervals (shaded area) are shown.

Table 7: **Model parameterisation (posterior parameters).** Notation, a brief description, and the values used for two health zones Bagata and Mosango. Values were estimated in Crump *et al.* (9).

| Notation | Description | Value (median and 95% CI) Mosango | Bagata | Unit |
|---|---|---|---|---|
| $R_0$ | Basic reproduction number (NGM approach) | 1.012 [1.007, 1.026] | 1.017 [1.010, 1.032] | - |
| $r$ | Relative bites taken on high-risk humans | 3.24 [1.68, 6.54] | 5.93 [3.22, 11.00] | - |
| $k_1$ | Proportion of low-risk people | 0.927 [0.798, 0.978] | 0.857 [0.723, 0.941] | - |
| $k_4$ | Proportion of high-risk people | $k_4 = 1 - k_1$ | | - |
| $\eta_H^{\text{post}}$ | Treatment rate from stage 1 (1998) | $1.06\times10^{-4}$ $[0.36, 2.53]\times10^{-4}$ | $2.25\times10^{-4}$ $[1.11, 4.51]\times10^{-4}$ | days$^{-1}$ |
| $\gamma_H^{\text{pre}}$ | Treatment/death rate from stage 2 (pre-1998) | $2.06\times10^{-3}$ $[0.83, 5.20]\times10^{-3}$ | $1.22\times10^{-3}$ $[0.461, 3.36]\times10^{-3}$ | days$^{-1}$ |
| $\gamma_H^{\text{post}}$ | Treatment/death rate from stage 2 (1998) | $2.54\times10^{-3}$ $[1.15, 6.17]\times10^{-3}$ | $1.33\times10^{-3}$ $[0.57, 3.58]\times10^{-3}$ | days$^{-1}$ |
| Spec | Active screening diagnostic specificity | 0.9992 [0.9987, 0.9997] | 0.9992 [0.9989, 0.9995] | - |
| $u$ | Proportion of passive cases reported | 0.329 [0.238, 0.433] | 0.291 [0.195, 0.402] | - |
| $\eta_{H\text{amp}}$ | Relative improvement in passive stage 1 detection rate | 1.046 [0.18, 3.58] | 3.46 [1.22, 7.64] | - |
| $\gamma_{H\text{amp}}$ | Relative improvement in treatment/death rate from stage 2 | 0.33 [0.04, 0.92] | 0.068 [0.002, 0.433] | - |
| $d_{\text{steep}}$ | Speed of improvement in passive detection rate | 1.04 [0.73, 1.39] | 1.06 [0.76, 1.39] | - |
| $d_{\text{change}}$ | Midpoint year for passive improvement | 2004.9 [2002.7, 2010.0] | 2011.1 [2007.9, 2013.2] | - |

### 3 Model comparison

#### 3.1 Differences between data extraction

Data used for fitting of Model W (2000–2016) were extracted from the WHO HAT Atlas by using geolocation information (where available) and following the detailed algorithm laid out in the Supplementary Information of Crump *et al.* (9). The data for screening in 2017 and 2018, used as part of model projections was extracted from an updated version of the HAT Atlas using health zone name information alone. These data have no impact on the fit of Model W.

Model S also used WHO HAT Atlas data (2000–2016) for fitting Mosango, and 2000–2018 for fitting Bagata. Screening coverage of the years 2017–2018 was also used in projections, however rather than geolocations, extraction was performed using health zone names. The comparison of data extracted using the two different methods is seen in Figure 7. It is noted that there are some marginal discrepancies in the two health zones selected, most noticeably in Mosango where the geolocation extraction yields slightly more cases in both active and passive detection for most of 2000–2010 compared to health zone name extraction.

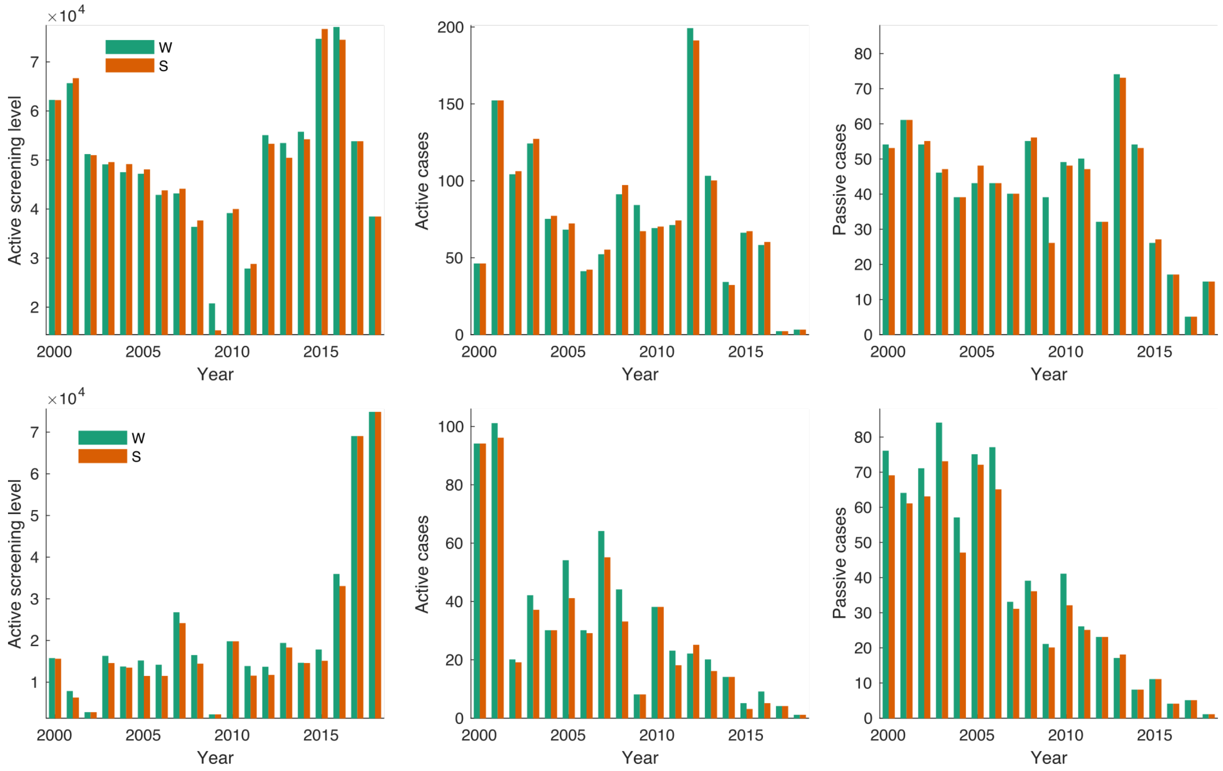


Figure 7: Comparison of data extracted for fitting of the two different models. Data used in Model W is shown in green, and data used in Model S is shown in orange. The top row shows data for Bagata health zone, and the bottom for Mosango.

#### 3.2 Model structure comparison

A comparison of the stuctures of the two models is shown in Table 8.

Table 8: **Model comparison**. The differences between the modelling assumptions of Model W and Model S.

| | **Model W** | **Model S** |
|---|---|---|
| **Risk structure in human populations** | Yes — high-/ low-risk structure. High-risk not participating in active screening and higher relative risk of tsetse bites. | Yes — high-/ low-risk structure. High-risk not participating in active screening and higher relative risk of tsetse bites. Human mobility between regions of low and high tsetse exposure. |
| **Model fitting** | Using fitting from deterministic model to 2000–2016 data. Adaptive MCMC method is described in Crump et al (9). | Using fitting from deterministic model to 2000–2016/2018 data (Mosango/Bagata health zone respectively). Adaptive MCMC method and output are found in SI Model S - model fit. |
| **Stochastic model for simulations** | Tau-leap method (one day time step). Tsetse with ODE equations. | Adaptive tau-leap method applied to the whole dynamical system. |
| **Improvements in passive surveillance** | Yes — in 1998 (following the introduction of the CATT test). Afterwards there is a gradual improvement using the equations described in Section 2.1. | Yes — Constant rates before 2000, improvement assumed to start afterwards and fitted following equations in section 1.1. |
| **Active screening** | Occurs once (pulsed) at the beginning of each year. | Continuous rate over each year. |
| **Imperfect specificity** | Fitted for each health zone independently, however median posterior values were both 99.92% in both Mosango and Bagata. Switch to perfect specificity in 2015 (Mosango) and 2018 (Bagata). | Fitted for each health zone independently, with median in Mosango of 99.86% and Bagata of 99.98%. Switch to perfect specificity in 2015 (Mosango) and 2018 (Bagata). |
| **Animal or asymptomatic reservoir** | None assumed. | None assumed. |

### 4 NTD-PRIME criteria

Table 9: **PRIME-NTD criteria fulfillment**. How this study has satisfied the five principles of the Neglected Tropical Diseases Modelling Consortium.

| The principle and what has been done to satisfy this principle? | Where in the manuscript is this described? |
|---|---|
| **1. Stakeholder engagement** | |
| The study was led by modellers with guidance from E Mwamba Miaka of the national sleeping sickness control programme in DRC (PNLTHA-DRC). PNLTHA-DRC have provided detailed knowledge of the data and information as to how the programme has changed over time. | Authorship list. |
| **2. Complete model documentation** | |
| The deterministic model code used for fitting Model W is available on OpenScienceFramework (OSF) at https://osf.io/ck3tr/. The models are fully described in the supplementary material. | OSF and Supplementary material. |
| **3. Complete description of data used** | |
| Our models are fitted to screening and case data from the WHO HAT Atlas for the selected health zones. The fitting process for Model W is described in detail in (9) and of Model S in the supplementary information. | Methods section of main manuscript, supplementary information and (9). |
| **4. Communicating uncertainty** | |
| Structural uncertainty: Model selection for Model W was completed in Rock et al. (2015), where plausible models were compared.<br>Model parameter uncertainty: Models S and W were fitted using MCMC methodology with data specific to the health zones considered. 200 parameter sets from the generated posteriors were used to compute the infection dynamics in this study.<br>Stochastic uncertainty: Models S and W derive expected infection dynamics from 200,000 stochastic realisations. The confidence interval of the mean is negligible due to the large number of simulations. | Methods and supplementary material. |
| **5. Testable model outcomes** | |
| We provide predicted future case reporting dynamics under a range of possible future scenarios which could be used to compare model outputs to observed data | Results section of main manuscript. |
