## Supplementary material for "Predicting the impact of COVID-19 interruptions on transmission of gambiense human African trypanosomiasis in two health zones of the Democratic Republic of Congo": Model S Fitting

### Supplementary information
### Model S - Fitting gHAT model to health zone data

This is a complimentary document to the main Supplementary Information material attached to the manuscript *Predicting the impact of COVID-19 interruptions on transmission of gambiense human African trypanosomiasis in two health zones of the Democratic Republic of Congo*.
This document presents additional information on Model S calibration to two health zones of the Democratic Republic of Congo, namely Bagata and Mosango health zones.

#### 1 Fitting procedure

Twelve parameters were fitted using annual case data from Mosango health zone for the period 2000–2016, and for Bagata for the time period 2000–2018. The model was first run to reach equilibrium prevalence of infection assuming only constant passive screening before 2000, when the fitting starts. The data that was fitted, distinguished reported cases from active screening and passive detection, and provides information on staging only after 2015. Fitting was performed via an adaptive Metropolis-Hastings Markov chain Monte Carlo (MCMC) approach, and used the following log-likelihood function:

$$\begin{aligned} LL(\theta|x) = & \log(P(x|\theta)) \\ \propto & \sum_{i=2000}^{2016 \text{ or } 2018} \Bigg( \log\left[\text{NegBin}\left(A_{d1}(i) + A_{d2}(i), A_{m1}(i) + A_{m2}(i), \kappa_{\text{AS}}\right)\right] \\ & + \log\left[\text{NegBin}\left(P_{d1}(i) + P_{d2}(i), P_{m1}(i) + P_{m2}(i), \kappa_{\text{PD}}\right)\right] \\ & + \log\left[\text{Bin}\left(P_{d1}(i), P_{d1}(i) + P_{d2}(i), \frac{P_{m1}(i)}{P_{m1}(i) + P_{m2}(i)}\right)\right] \\ & + \log\left[\text{Bin}\left(A_{d1}(i), A_{d1}(i) + A_{d2}(i), \frac{A_{m1}(i)}{A_{m1}(i) + A_{m2}(i)}\right)\right] \Bigg) \end{aligned}$$

The first two terms in the inner sum represent the total number of active and passive cases in the data, respectively, modeled as a negative binomial with mean equal to the number of cases from the differential equation model. The second two terms in the inner sum represent the proportion of cases in stage 1, modeled as a binomial in which the probability of stage 1 is the proportion from the differential equation model (and the number of trials is from the data of total number of cases). Since we only have staged data for after 2015, those terms only contribute to the log likelihood in those years.

To sample from the posterior distribution, determined by the likelihood function and the prior distributions, we used an adaptive Metropolis-Hastings MCMC algorithm (unpublished, Spencer). For the sampling, we ran two independent chains of the algorithm to corroborate convergence. We used a burn-in period of 2000 steps and then ran the chain for 20,000 steps, which was thinned to every other sample. For the proposal distribution, we used a multivariate Normal distribution (truncated with the bounds given in Table 1), with a covariance matrix that adapts to predict the shape and scale of the posterior distribution as the algorithm proceeds. The adaptation improves the efficiency of proposing new samples so that there are neither excessive rejections nor acceptances in the algorithm. Finally, to improve mixing further, we used the two covariance matrices from this first set of runs in a second round of two independent chains with the same burn-in and sampling strategy. With this second set, we visually checked that there was good mixing. The parameters used in the forward projections are based on the first chain of this second set of runs.

#### 1.1 Fixed parameters, priors and posterior distributions

Descriptions and values of all fixed parameters are given in Table 2 of the main Supplementary Information file, and descriptions and prior distributions for all fitted parameters are given in Table 1 and Table 2 respectively.

| **Parameter** | **Unit** | **Prior Distribution and Bounds Mosango** | **Bagata** |
|---|---|---|---|
| $\kappa$ | - | Beta$(1, 500)$ in $[0, 1]$ | Beta$(1, 500)$ in $[0, 0.63]$ |
| log(VHL) | - | N$(1.1, 0.0025)$ in $[0, 10]$ | same |
| log($c_1$) | - | Gamma$(1, 1)$ in $[0, 10]$ | same |
| logit(spec) | - | Unif$[6.212606, 9.21024]$ | same |
| $r1_{\text{const}}$ | day$^{-1}$ | Unif$[0, 0.001]$ | same |
| log($c_2$) | - | Gamma$(1, 1)$ in $[0, 10]$ | same |
| $\Delta r_1$ | year$^{-1}$ | Unif$[0, 2.5]$ | same |
| $\Delta r_2$ | year$^{-1}$ | Unif$[0, 2.5]$ | same |
| $x_0$ | - | Gamma$(10, 0.27)$ in $[0, 17]$ | Unif$[0, 19]$ |
| $\alpha_{\text{pd}}$ | - | Unif$[0.1, 5]$ | same |
| $\kappa_{\text{as}}$ | - | Gamma$(23.5, 3)$ | same |
| $\kappa_{\text{pd}}$ | - | Gamma$(23.5, 3)$ | same |

Table 1: Priors for fitted parameters for Model S. Non-uniform priors were additionally truncated with values given in brackets. Gamma priors are written with arguments of shape and scale.

Table 2: **Model parameterisation (posteriors of fitted parameters).** Notation, a brief description, and representative percentiles of the posterior distributions for fitted parameters. Here logarithm always refers to the natural logarithm.

| Notation | Description | Posterior (median [95% CI]) | |
|---|---|---|---|
| | | **Bagata** | **Mosango** |
| $\kappa$ | Ratio of humans in the high- to low-exposure environment | 7.03 $\times 10^{-4}$ [ 1.27 , 20.5]$\times 10^{-4}$ | 4.52 $\times 10^{-3}$ [3.41, 5.98]$\times 10^{-3}$ |
| log(VHL) | Log ratio of vectors to humans in low-exposure environment | 1.359 [1.312, 1.386] | 1.180 [1.159, 1.207] |
| log($c_1$) | Log ratio of the ratio of vectors to humans in the high exposure environment to the ratio of vectors to humans in the low exposure environment | 8.58 $\times 10^{-2}$ [0.43, 89.29]$\times 10^{-2}$ | 2.44$\times 10^{-2}$ [0.099, 10.26]$\times 10^{-2}$ |
| logit(spec) | Diagnostic specificity (active screening) | 8.815 [8.382, 9.183] | 6.571 [6.331, 6.807] |
| $r1_{\text{const}}$ | Daily passive detection rate for stage 1 (pre-2000) | 3.33$\times 10^{-6}$ [0.28, 12.6]$\times 10^{-6}$ | 1.33$\times 10^{-5}$ [0.042, 5.35]$\times 10^{-5}$ |
| log($c_2$) | Log ratio of passive detection for stage 2 to stage 1 (pre-2000) | 0.331 [0.015, 0.953] | 0.466 [.0134, 2.163] |
| $\Delta r_1$ | Amount passive detection in stage 1 improves | 0.519 [0.223, 0.106] | 0.289 [0.207, 0.404] |
| $\Delta r_2$ | Amount passive detection in stage 2 improves (in addition to improvement of stage 1) | 0.704 [0.146, 1.766] | 0.554 [0.0838, 1.484] |
| $x_0$ | Turning point (years since 1999) for logistic improvement in passive detection | 13.75 [9.63, 17.68] | 2.63 [1.50, 4.33] |
| $\alpha_{\text{pd}}$ | Steepness in logistic improvement of passive detection | 0.327 [0.280, 0.390] | 0.405 [0.260, 0.623] |
| $\kappa_{\text{as}}$ | Overdispersion parameter (active screening) | 2.01 [1.45, 2.93] | 35.98 [20.88, 59.29] |
| $\kappa_{\text{pd}}$ | Overdispersion parameter (passive detection) | 48.96 [29.81, 76.06] | 66.06 [42.01, 97.58] |

#### 1.2 MCMC outputs

The MCMC outputs shown in Figures 1 and 2 correspond to 10,000 post burn-in samples.

##### 1.2.1 Posterior densities

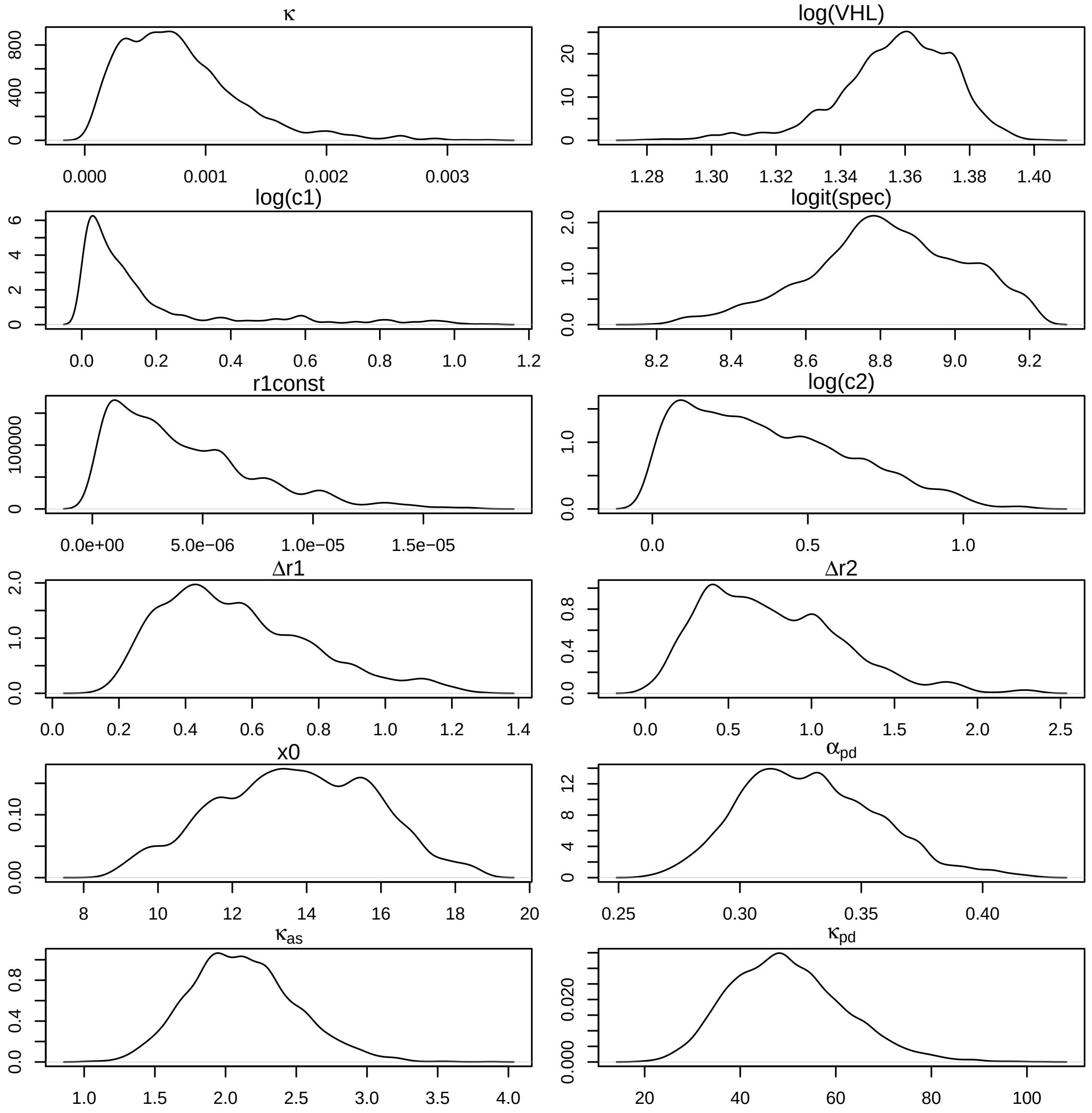


Figure 1: Posterior density of fitted parameters for Bagata health zone.

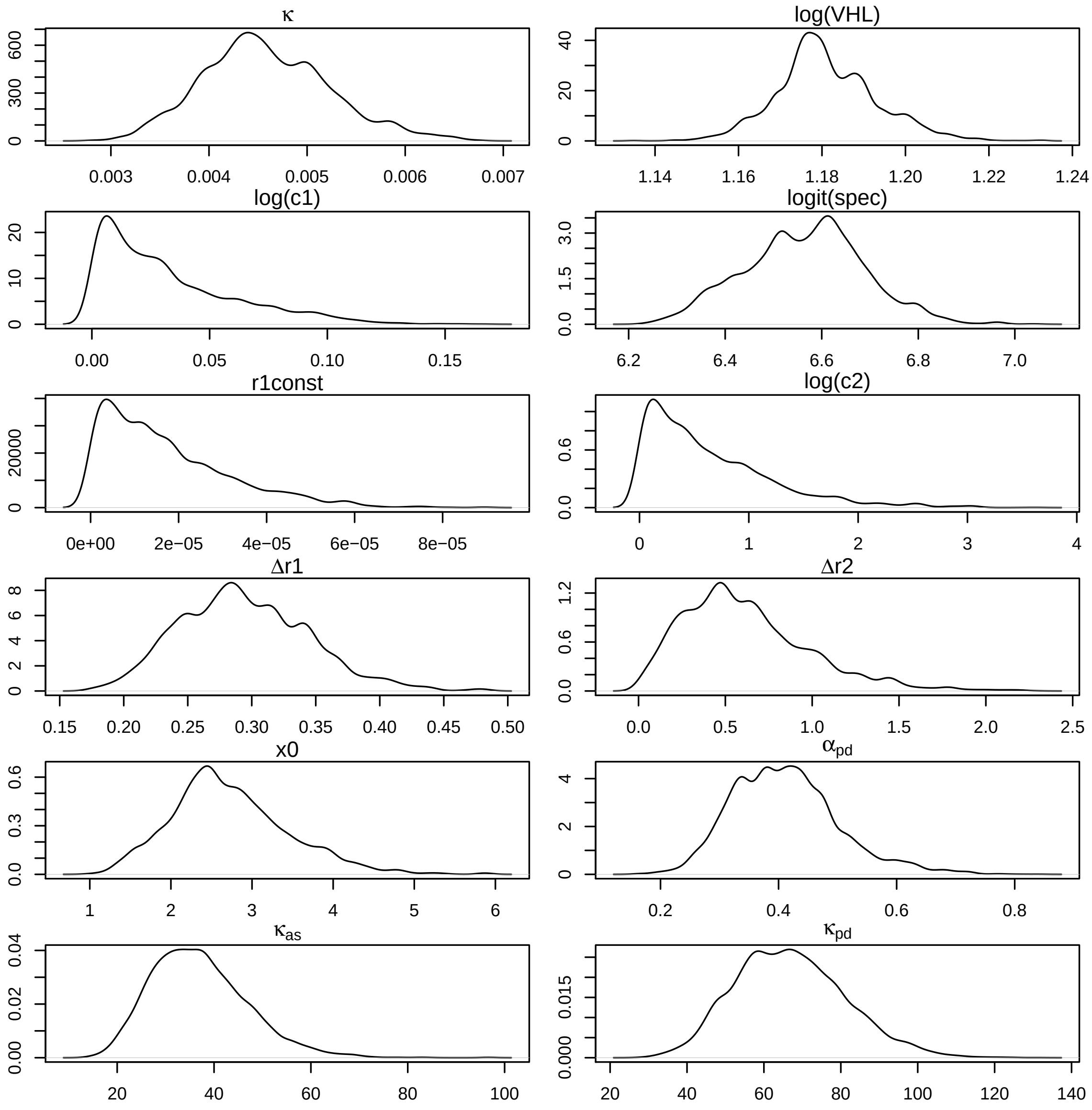


Figure 2: Posterior density of fitted parameters for Mosango health zone.

##### 1.2.2 Model fit to reported case data

It is worth noting that while the model could capture the number of reported cases through active screening all along the data period for Mosango health zone (Figure 3), it overestimated the active reported cases for Bagata health zone in the first years (Figure 4). One of the model assumptions is that there was no active screening before 2000 because we had no data before 2000; and it is possible that some active screening had been conducted in Bagata before 2000, that would have reduced prevalence to levels below what the model predicted. However, after a few years of active screening, the model is able to capture the number of reported cased from active screening (see Figure 5).

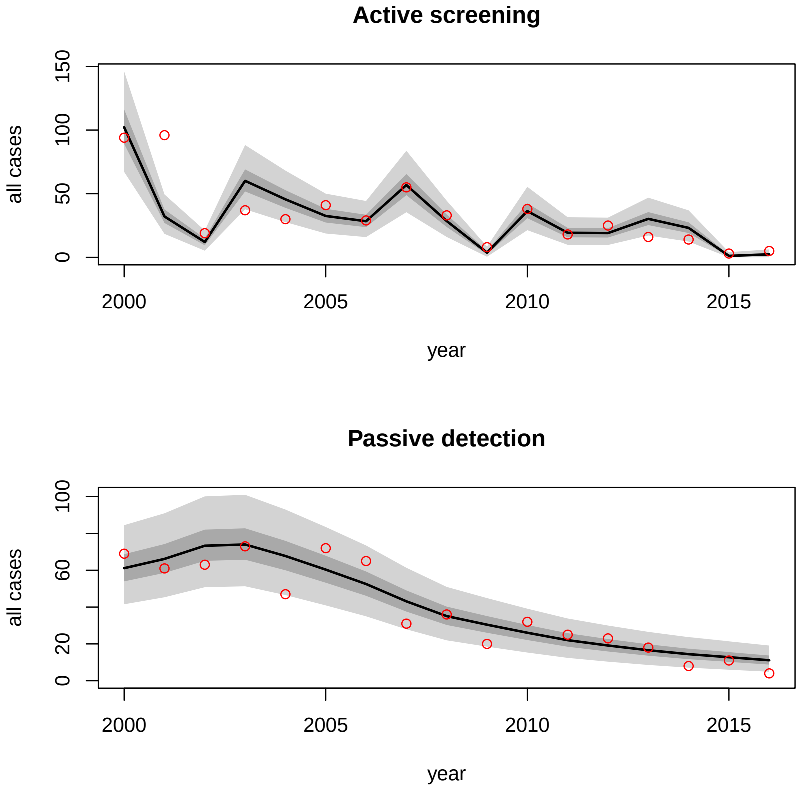


Figure 3: Model fit to reported cased data for Mosango health zone.

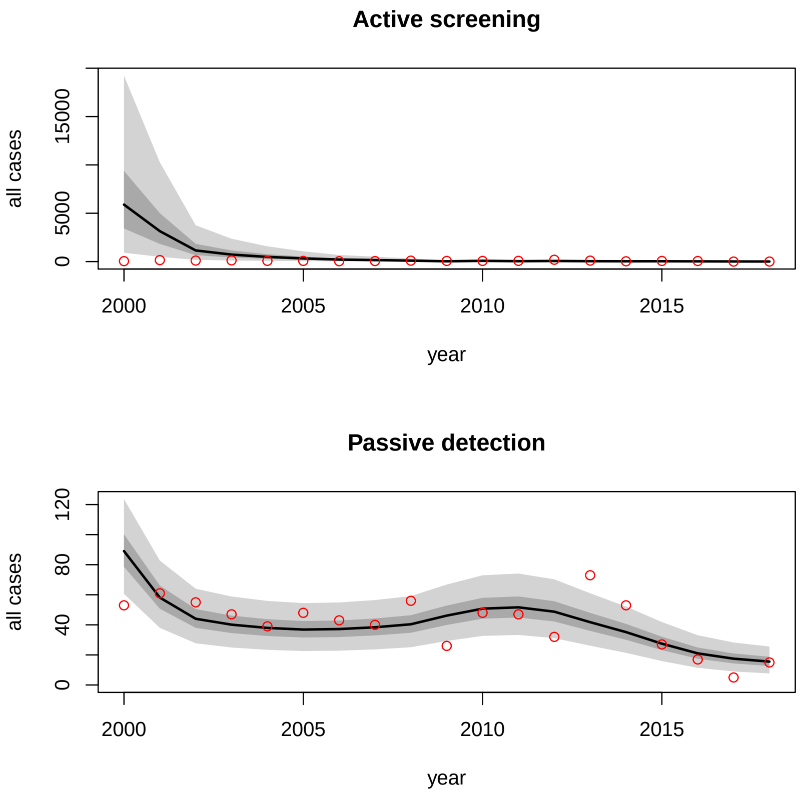


Figure 4: Model fit to reported cased data for Bagata health zone.

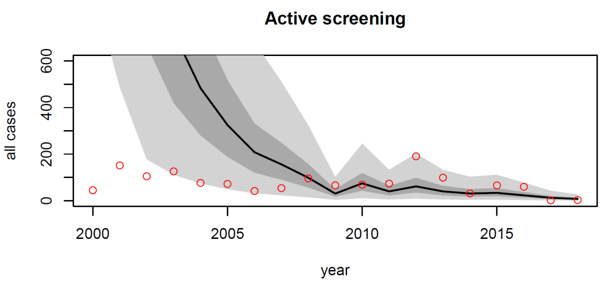


Figure 5: Reduced scale showing model fit to active reported data for Bagata health zone.
